# Hypertension disease natural history and healthcare utilization when treatment guidelines do not account for race

**DOI:** 10.64898/2026.10.05.26364804

**Authors:** Marika M. Cusick, Malcolm Barrett, Fernando Alarid-Escudero, Jeremy D. Goldhaber-Fiebert, Sherri Rose

## Abstract

**Objective:** To estimate the impact of removing race-adjusted antihypertensive treatment recommendations for Black adults on health outcomes, healthcare utilization, and racial/ethnic disparities.

**Study Setting and Design:** We conducted a decision-analytic modeling study using a discrete-time state-transition microsimulation of hypertension natural history and health system utilization. We evaluated the effect of removing race-adjusted antihypertensive treatment recommendations under two scenarios: one assuming racial heterogeneity in treatment effectiveness, reflecting evidence behind prior race-specific guidelines, and the other assuming no racial heterogeneity. Main outcomes included life expectancy, cumulative incidence and duration of hypertension, and blood pressure control with antihypertensive treatment.

**Data Sources and Analytic Sample:** The microsimulation simulated lifetime trajectories among 100,000 non-Hispanic Black and non-Hispanic white 40-year-old adults. Model parameters were derived from multiple data sources, including the 2013-2018 National Health and Nutrition Examination Survey, the American Board of Family Medicine PRIME primary care electronic health record registry from 2018 and 2024, 2021 U.S. life tables, and published literature.

**Principal Findings:** Under assumptions of heterogeneous treatment effectiveness by race, guidelines without race adjustment decreased life expectancy among non-Hispanic Black adults by 0.4 years, increasing the life expectancy gap with non-Hispanic white adults from 4.1 to 4.5 years. Hypertension control among non-Hispanic Black adults also worsened. Years with uncontrolled hypertension increased by 0.8 years, widening the disparity with non-Hispanic white adults from 1.7 to 2.5 years. Assuming no racial heterogeneity, these effects were diminished: life expectancy decreased by 0.1 years and years with uncontrolled hypertension increased by 0.2 years, less than one-half and one-quarter of the effects observed under assumptions of heterogeneous treatment effectiveness by race.

**Conclusions:** The potential effects of removing race-adjusted antihypertensive treatment guidelines depended on assumptions of racial heterogeneity in treatment effectiveness. Our findings highlight the importance of understanding drivers of previously observed racial differences when considering modifying antihypertensive treatment guidelines.

## 1 Introduction

Hypertension affects nearly half of American adults and is a major driver of cardiovascular mortality in the U.S.^1^ The burden of hypertension is not shared equally across the population. Non-Hispanic Black adults have a higher prevalence of hypertension compared with non-Hispanic white, non-Hispanic Asian, and Hispanic adults.^2,3^ Furthermore, only one in four adults with hypertension achieve recommended blood pressure targets,^4^ and rates of blood pressure control are lower among non-Hispanic Black, Hispanic, and non-Hispanic Asian adults compared with non-Hispanic white adults.^5^ Previous research suggests that variation in hypertension control is shaped by socioeconomic factors, including income, insurance status, or educational attainment.^4,5^

Most adults with hypertension are advised to make lifestyle modifications and initiate antihypertensive treatment to lower blood pressure and reduce the risk of cardiovascular events.^4,6^ The Antihypertensive and Lipid-Lowering Treatment to Prevent Heart Attack Trial (ALLHAT), conducted between 1994 and 2002, compared antihypertensive medication classes in preventing major cardiovascular events. Subgroup analyses from ALLHAT revealed heterogeneity in antihypertensive treatment effectiveness between Black and non-Black adults.^7–9^ These findings informed race-specific pharmacologic therapy recommendations for Black adults in major clinical practice guidelines, such as the 2014 Eighth Joint National Committee, 2017 American Heart Association (AHA)/American College of Cardiology (ACC), and 2018 European Society of Cardiology guidelines.^6,10,11^

However, the inclusion of race in clinical algorithms and practice guidelines has faced growing criticism for propagating racial bias in clinical decision-making and exacerbating health disparities.^12–14^ In response, national organizations have removed race or ethnicity-based adjustments from some clinical equations.^15–19^ Recently, the 2025 AHA/ACC guidelines on the prevention, detection, evaluation, and management of high blood pressure in adults removed race-specific recommendations for antihypertensive treatment.^20^

The implications of removing race-adjusted antihypertensive treatment recommendations for health outcomes and racial/ethnic disparities remain unclear, in particular given uncertainty in the existence of racial heterogeneity in treatment effectiveness. Rigorous evaluations of clinical guideline changes are critical to assess their potential impact and minimize unintended harm.^21^ In this study, we developed a microsimulation model to estimate the impact of removing race-adjusted recommendations on health outcomes, healthcare utilization, and racial/ethnic disparities between non-Hispanic Black and non-Hispanic white adults. To reflect the uncertainty of racial heterogeneity in treatment effectiveness, we considered two scenarios: one assuming race-specific treatment effects and another assuming equal effectiveness across racial groups.

## 2 Methods

### 2.1 Microsimulation model overview

We conducted a decision-analytic modeling study using a discrete-time state transition microsimulation model. The microsimulation model had two interacting processes: disease natural history and health system utilization (Figure 1). The disease natural history process simulated individual systolic and diastolic blood pressure trajectories and progression to death. The health system process tracked individuals through three stages: in the health system (with routine place for healthcare), detected and treated for hypertension, and discontinued treatment.

**Figure 1:**
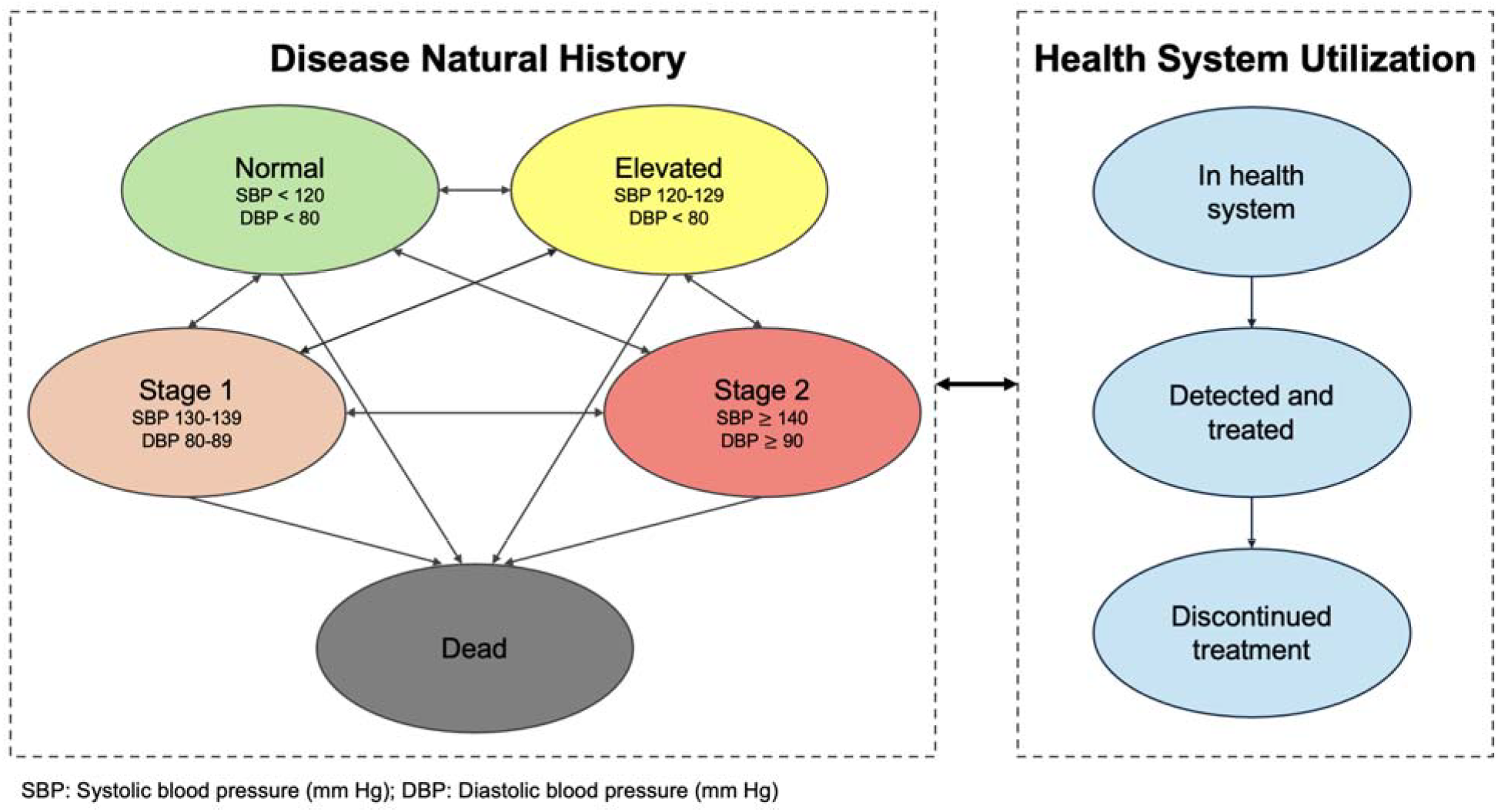
Model diagram of state-transition microsimulation model of hypertension disease natural history and health system utilization

### 2.2 Data sources

Multiple data sources were used to parameterize our microsimulation model (Table S1). To obtain a simulation cohort representative of U.S. adults, we relied on survey data from the 2013-2014, 2015-2016, and 2017-2018 years of the National Health and Nutrition Examination Survey (NHANES).^22^ We sampled one million blood pressure measurements between January 2018 and June 2024 from the American Board of Family Medicine PRIME primary care electronic health record registry to estimate model transitions in hypertension natural history and health system utilization.^23,24^ Baseline mortality rates were derived from 2021 age-, sex-, race-, and ethnicity-specific life tables.^25^ Remaining model parameters, including the effectiveness of antihypertensive treatments in lowering blood pressure, antihypertensive monotherapy utilization, and treatment discontinuation rates, were obtained from published literature as cited.

### 2.3 Simulation cohort

Our simulation cohort consisted of 100,000 synthetic 40-year-old non-Hispanic Black and non-Hispanic white adults. We sampled synthetic individuals using a multivariate copula model fit to NHANES data (Section S 6.2). Synthetic individuals differed by race/ethnicity, sex, initial systolic and diastolic blood pressure values, and blood pressure measurement frequencies.

### 2.4 Disease natural history

The microsimulation model simulated lifetime systolic and diastolic blood pressure trajectories. Blood pressure values were categorized into hypertension stages using standard blood pressure cut-offs: normal (systolic <120 mm Hg and diastolic <80 mm Hg), elevated (systolic 120-129 mm Hg and diastolic <80 mm Hg), stage 1 (systolic 130-139 mm Hg or diastolic 80-89 mm Hg), and stage 2 (systolic ≥ 140 mm Hg or diastolic ≥ 90 mm Hg).^26^ To allow for individual variation in blood pressure, we fit quantile regressions to predict monthly systolic and diastolic blood pressure values using the PRIME registry (Section S 6.3). Assigned quantiles for blood pressure prediction were updated every five years according to an exponential distribution, while preserving correlations between systolic and diastolic blood pressure values observed in the PRIME registry (Section S 6.4). Treatment initiation lowered blood pressure according to treatment effects observed in the ALLHAT clinical trial, further described in Section 2.6. Individuals who discontinued treatment returned to blood pressure trajectories before treatment exposure. Mortality rates varied by age, sex, race/ethnicity, and blood pressure. We used age-sex-race/ethnicity-specific mortality rates from the 2021 U.S. life tables.^25^ Individuals with elevated blood pressure experienced increased mortality risk (Section S 6.5).^27^

### 2.5 Health system utilization

All synthetic individuals were initialized as “in the health system” state (Figure 1). Treatment initiation was simulated in two steps. First, individual blood pressure measurement intervals ranged from monthly to annually according to their age, sex, and prior blood pressure values. Second, at each blood pressure measurement, the probability of initiating antihypertensive therapy was predicted as a function of age, sex, and systolic and diastolic blood pressure values (Section S 6.6). Given limitations of electronic health record data in measuring treatment discontinuation, treatment class-specific discontinuation rates were obtained from the ALLHAT trial.^28^ Our model assumed that individuals who discontinued treatment did not reinitiate treatment.

### 2.6 Interventions

We compared the 2025 AHA/ACC guideline, which removed race-specific antihypertensive treatment recommendations, with continued use of the 2017 AHA/ACC race-adjusted recommendations. Our analysis focused on first-line monotherapy with thiazide diuretics or angiotensin-converting enzyme (ACE) inhibitors among individuals with stage 1 or 2 hypertension. The 2017 AHA/ACC guidelines recommended thiazide diuretics or ACE inhibitors as first-line antihypertensive therapy. However, these guidelines also included a special recommendation for Black adults, removing ACE inhibitors as a first-line therapy for this racial group.^6^ In simulating race-adjusted guidelines, non-Hispanic Black adults were prescribed thiazide diuretics only, whereas non-Hispanic white adults were prescribed both thiazide diuretics and ACE inhibitors.^29^ The 2025 AHA/ACC guideline no longer includes a special recommendation by race. Our simulation assumed that prescribing patterns for thiazide diuretics and ACE inhibitors did not differ by race.

Guideline changes were evaluated under two treatment effect scenarios: 1) heterogeneity in treatment effectiveness by race and 2) no heterogeneity in treatment effectiveness by race. In the first scenario, we used ALLHAT subgroup analyses that demonstrated treatment heterogeneity by race: ACE inhibitors were inferior to thiazide diuretics in blood pressure control and cardiovascular disease prevention among Black adults.^7^ Blood pressure reductions and discontinuation rates differed by race in the simulation. In the second scenario, treatment effects and discontinuation rates were based on estimates from the overall ALLHAT trial population and were applied equally across racial/ethnic groups.^28^ Blood pressure reductions varied by years on treatment through year four of ALLHAT follow-up (Table 1). Simulated individuals who remained on treatment after year four retained the year-four effect.

**Table 1:** Absolute reductions in blood pressure (systolic/diastolic, mm Hg) by antihypertensive treatment type and treatment effect heterogeneity assumption.

| Antihypertensive Treatment | Year | Heterogeneous Treatment Effects |  | No Heterogeneous Treatment Effects |  |
| --- | --- | --- | --- | --- | --- |
|  |  | Black | Non-Black | Black | Non-Black |
| Thiazide diuretic | 1 | 7.7/3.9 | 9.8/4.7 | 9.3/4.7 | 9.3/4.7 |
|  | 2 | 8.6/5.0 | 10.6/5.8 | 10.3/5.7 | 10.3/5.7 |
|  | 4 | 10.5/6.6 | 12.3/7.6 | 12.3/7.5 | 12.3/7.5 |
| ACE inhibitor | 1 | 2.5/2.3 | 8.1/4.9 | 6.1/4.2 | 6.1/4.2 |
|  | 2 | 3.4/3.4 | 9.5/6.1 | 8.0/5.5 | 8.0/5.5 |
|  | 4 | 6.8/5.6 | 12.0/8.0 | 10.9/7.5 | 10.9/7.5 |

ACE inhibitor: angiotensin-converting enzyme inhibitor

### 2.7 Outcomes and analysis

To validate the hypertension disease natural history process, we simulated a hypothetical cohort of 10,000 individuals from the PRIME registry before antihypertensive treatment exposure (Section S 6.7). Our main outcomes included life expectancy, years spent with stage 1 or 2 hypertension, cumulative incidence and duration of antihypertensive treatment, and years with uncontrolled hypertension. We defined uncontrolled hypertension as having stage 1 or stage 2 blood pressure values while receiving treatment. We reported mean estimates and 95% confidence intervals with Monte Carlo standard errors (MCSEs) for all outcomes.

### 2.8 Secondary analysis

In a secondary analysis, we leveraged the previously published social factors framework, a decision-modeling approach for health policy analyses when outcomes are influenced by both disease and social processes.^30^ Two social factors were additionally modeled in our microsimulation model: health insurance status and census tract-level social deprivation index (SDI) (Section S 6.8, Figure S6, Table S9). Census tract-level SDI served as a proxy for socioeconomic status and captured geographic variation in hypertension prevalence, awareness, and control.^4,5^ The updated model included a fourth health system stage, out of the health system, representing individuals without a routine place for healthcare. At model initiation, synthetic individuals were assigned a census tract-level SDI, insurance status, and an initial health system state (Section S 6.2, Section S 6.9).^31^. After age 65, all simulated individuals were assumed to have insurance through Medicare coverage.

To incorporate the impact of the social factors on disease natural history, we modified several model parameters and transition probability dependencies. Baseline mortality rates were further adjusted by insurance status.^32–34^ Having no health insurance increased baseline mortality rates, assuming a hazard ratio for increased mortality risk among those uninsured (Section S 6.10). We predicted systolic and diastolic blood pressure trajectories as a function of age, sex, and census tract-level SDI (Section S 6.3, Section S 6.4).

Insurance status and census tract-level SDI also influenced health system utilization. Uninsured individuals were less likely to enter the health system and had elevated blood pressures compared to insured individuals, as observed in NHANES (Section S 6.11, Section S 6.12). Treatment initiation rates varied by census tract-level SDI. We predicted blood pressure measurement frequencies and antihypertensive treatment probabilities as a function of age, sex, systolic and diastolic blood pressure values, and census tract-level SDI (Section S 6.6). Finally, uninsured individuals were assumed to discontinue treatment at faster rates compared to insured individuals.^35^

## 3 Results

### 3.1 Cohort characteristics

The simulation cohort included 100,000 adults aged 40 years, evenly divided between non-Hispanic Black and non-Hispanic white individuals. At age 40, non-Hispanic Black adults had a higher proportion of individuals in hypertension stages 1 (36% vs. 34%) and stage 2 (19% vs. 14%) compared to non-Hispanic white adults (Table 2). In model validation, simulation model outputs were concordant with age-specific hypertension prevalence estimates in the PRIME registry (Table S8, Figure S4).

**Table 2:** Demographic and clinical characteristics of simulation cohort by race.

| Characteristic |  | Non-Hispanic Black (N=50,000) | Non-Hispanic white (N=50,000) |
| --- | --- | --- | --- |
| Female (%) |  | 52% | 51% |
| Hypertension stage (%) | Normal | 29% | 38% |
|  | Elevated | 15% | 15% |
|  | Stage 1 | 36% | 34% |
|  | Stage 2 | 19% | 14% |
| Systolic blood pressure (mean [SD]) |  | 124.1 [13.0] | 121.0 [12.7] |
| Diastolic blood pressure (mean [SD]) |  | 78.5 [9.1] | 77.0 [9.1] |

SD: Standard deviation

### 3.2 Heterogeneity in treatment effectiveness by race

Implementing antihypertensive guidelines without race adjustment shifted prescribing patterns for non-Hispanic Black adults, increasing the cumulative incidence of ACE inhibitor use from 0% to 46% and reducing thiazide diuretic use from 76% to 30% (Table S10). Prescribing patterns for non-Hispanic white adults were unchanged, with the cumulative incidence of ACE inhibitor and thiazide diuretic use at 45% and 30%, respectively. Because ACE inhibitors produced smaller blood pressure reductions and higher discontinuation rates among non-Hispanic Black adults, this prescribing shift had adverse consequences for health outcomes, healthcare utilization, and racial/ethnic disparities.

#### 3.2.1 Health outcomes

The cumulative incidence of stage 1 or 2 hypertension was unchanged at 90% under both guideline versions (Table S11). Implementing guidelines that no longer account for race increased time spent with hypertension for non-Hispanic Black adults. Years with stage 1 or 2 hypertension increased from 15.1 years (95% CI: 15.0, 15.2) under race-adjusted guidelines to 16.4 years (95% CI: 16.2, 16.5) (Figure 2 (a)). Life expectancy among non-Hispanic Black adults also decreased from 36.0 years (95% CI: 35.9, 36.1) to 35.7 years (95% CI: 35.5, 35.8). This decline widened the life expectancy gap with non-Hispanic white adults from 4.1 years to 4.5 years (Table 3, Figure 2 (b)).

**Table 3:** Main outcomes for non-Hispanic Black and non-Hispanic white adults comparing antihypertensive treatment guidelines with and without race adjustment.

| Outcome | Group | Heterogeneous Treatment Effects |  | No Heterogeneous Treatment Effects |  |
| --- | --- | --- | --- | --- | --- |
|  |  | Race Adjustment | No Race Adjustment | Race Adjustment | No Race Adjustment |
| Years spent in stage 1 or 2 hypertension | Non-Hispanic Black | 15.1 [0.1] | 16.4 [0.1] | 14.3 [0.1] | 14.8 [0.1] |
|  | Non-Hispanic white | 14.1 [0.1] | 14.1 [0.1] | 14.4 [0.1] | 14.4 [0.1] |
|  | Difference | -1.0 [0.1] | -2.3 [0.1] | 0.1 [0.1] | -0.4 [0.1] |
| Years on antihypertensive treatment | Non-Hispanic Black | 22.1 [0.1] | 20.4 [0.1] | 22.3 [0.1] | 21.4 [0.1] |
|  | Non-Hispanic white | 22.7 [0.1] | 22.7 [0.1] | 22.4 [0.1] | 22.4 [0.1] |
|  | Difference | 0.6 [0.1] | 2.3 [0.1] | 0.0 [0.1] | 1.0 [0.1] |
| Years with uncontrolled hypertension | Non-Hispanic Black | 9.0 [0.1] | 9.8 [0.1] | 8.0 [<0.01] | 8.2 [<0.01] |
|  | Non-Hispanic white | 7.3 [<0.01] | 7.3 [<0.01] | 7.6 [<0.01] | 7.6 [<0.01] |
|  | Difference | -1.7 [0.1] | -2.5 [0.1] | -0.4 [0.1] | -0.6 [0.1] |
| Life expectancy | Non-Hispanic Black | 36.0 [0.1] | 35.7 [0.1] | 36.2 [0.1] | 36.1 [0.1] |
|  | Non-Hispanic white | 40.1 [0.1] | 40.1 [0.1] | 40.0 [0.1] | 40.0 [0.1] |
|  | Difference | 4.1 [0.1] | 4.5 [0.1] | 3.8 [0.1] | 4.0 [0.1] |

Mean [Monte Carlo standard error]

**Figure 2:**
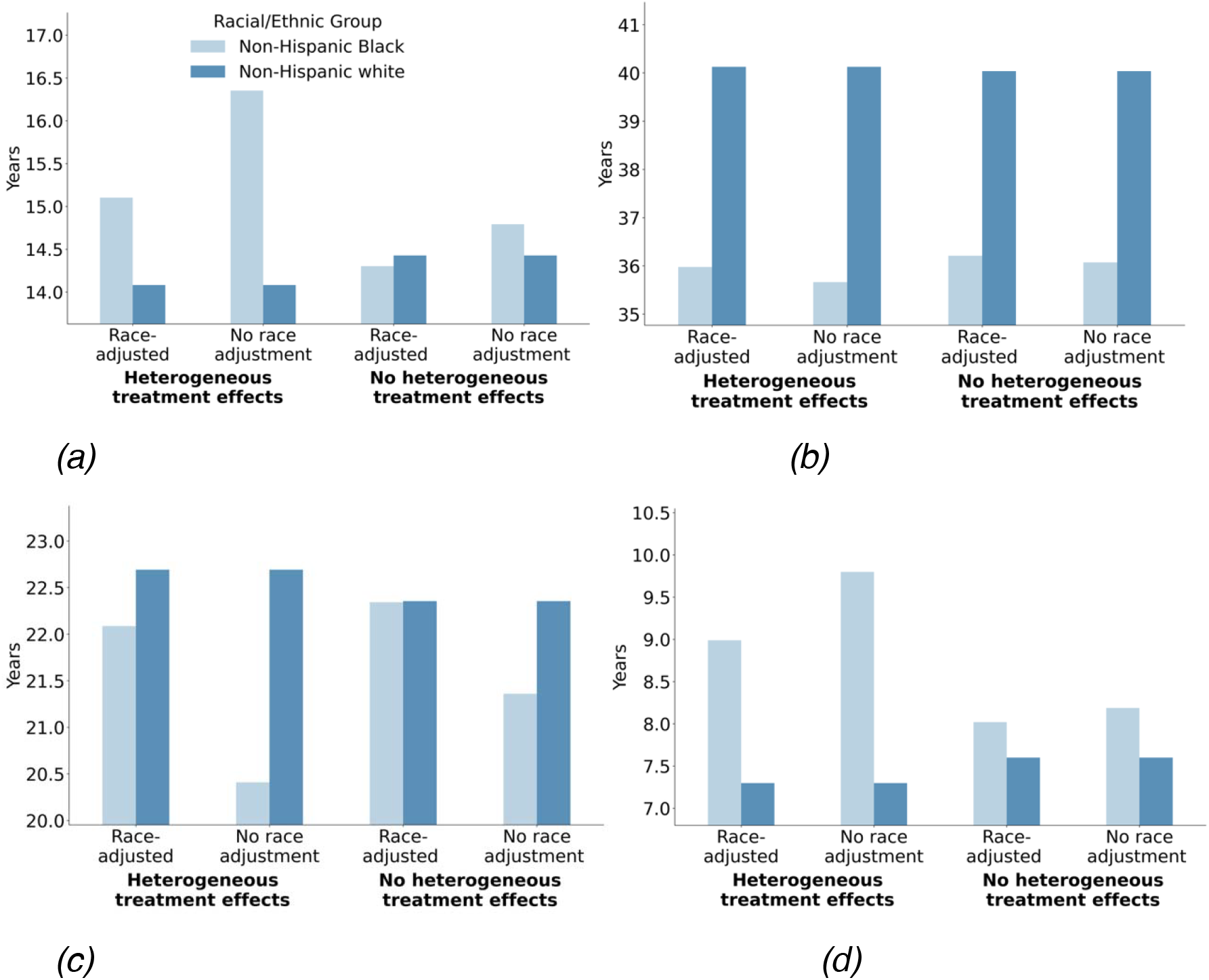
Comparing antihypertensive treatment guidelines with and without race adjustment for non-Hispanic Black and non-Hispanic white adults. Outcomes: (a) years with stage 1 or 2 hypertension, (b) life expectancy, (c) years on antihypertensive treatment, (d) years with uncontrolled hypertension

#### 3.2.2 Health system utilization

Treatment initiation was similar across groups, with the cumulative incidence of antihypertensive treatment of 76% in the overall cohort and across both racial groups (Table S11). The shift towards ACE inhibitors reduced the average duration of antihypertensive treatment among non-Hispanic Black adults from 22.1 years (95% CI: 22.0, 22.2) to 20.4 years (95% CI: 20.3, 20.5). As a result, the treatment duration gap with non-Hispanic white adults widened from 0.6 years to 2.3 years (Figure 2 (c)). Hypertension control also worsened. Among non-Hispanic Black adults, years with uncontrolled hypertension increased from 9.0 years (95% CI: 8.9, 9.1) to 9.8 years (95% CI: 9.7, 9.9) (Figure 2 (d)). The fraction of treatment time with uncontrolled hypertension rose from 41% to 49%, compared with 32% among non-Hispanic white adults.

### 3.3 No heterogeneity in treatment effectiveness by race

Under assumptions of no racial heterogeneity, the prescribing shift towards ACE inhibitors still worsened outcomes for non-Hispanic Black adults because ACE inhibitors were less effective at reducing blood pressure than thiazide diuretics overall. However, the magnitude of effects was smaller than in the heterogeneous effects scenario (Figure 2).

#### 3.3.1 Health outcomes

Guidelines without race adjustment increased years spent in stage 1 or 2 hypertension for non-Hispanic Black adults from 14.3 years (95% CI: 14.2, 14.4) to 14.8 years (95% CI: 14.7, 14.9), a 0.5-year increase compared to 1.3 years in the first scenario (Table 3, Figure 2). The effect on life expectancy among non-Hispanic Black adults was also smaller, decreasing from 36.2 years (95% CI: 36.1, 36.3) to 36.1 years (95% CI: 35.9, 36.2). This 0.1-year widening was less than half that observed in the first scenario (Table 3, Figure 2).

#### 3.3.2 Healthcare utilization

Healthcare utilization patterns were similar to the first scenario, though changes were smaller in magnitude. Average treatment duration among non-Hispanic Black adults decreased by 1.0 years, from 22.3 years (95% CI: 22.2, 22.5) to 21.4 years (95% CI: 21.2, 21.5), a smaller decline than the 1.7-year decrease in the first scenario (Table 3). Years with uncontrolled hypertension went from 8.0 years (95% CI: 7.9, 8.1) to 8.2 years (95% CI: 8.1, 8.3). As a result, the fraction of treatment time with uncontrolled hypertension rose by 3 percentage points, compared to 8 percentage points in the first scenario.

### 3.4 Secondary analysis

Insurance coverage was higher among non-Hispanic white 40-year-old adults (86%) than among non-Hispanic Black 40-year-old adults (79%). Mean census tract-level SDI was also higher for non-Hispanic Black adults (18.1) compared to non-Hispanic white adults (14.6), reflecting greater neighborhood disadvantage (Table S12). Incorporating census tract-level SDI and insurance coverage reduced overall treatment access. The overall incidence of treatment dropped from 76% in the main analysis to 69%. While treatment incidence did not differ by race, it differed by insurance status (Table S13). Among those without insurance, 58% and 57% of non-Hispanic Black and non-Hispanic white adults initiated treatment, compared to 71% and 70% of insured non-Hispanic Black and non-Hispanic white adults (Figure S10).

#### 3.4.1 Heterogeneity in treatment effectiveness by race

Under guidelines without race adjustment, life expectancy among non-Hispanic Black adults decreased by 0.3 years, from 35.5 years (95% CI: 35.3, 35.6) to 35.2 years (95% CI: 35.0, 35.3), although with overlapping confidence intervals. This effect was similar to the 0.4-year effect observed in the main analysis (Table S14, Figure S11). Because insured non-Hispanic Black adults had greater treatment access, they were more likely to be affected by the prescribing shift towards ACE inhibitors. The average decline in life expectancy was 0.3 years for non-Hispanic Black adults with insurance compared to 0.2 years for those without insurance (Table S15). We observed a similar pattern in hypertension control. Years spent with uncontrolled hypertension increased by 0.8 years and 0.2 years among non-Hispanic Black adults with and without insurance, respectively.

#### 3.4.2 No heterogeneity in treatment effectiveness by race

As in the main analysis, the effects of guidelines without race adjustment were smaller under the no treatment heterogeneity scenario. The decline in life expectancy for non-Hispanic Black adults was the same as the 0.1-year widening in the main analysis (Table S14, Figure S11). In this scenario, we observed no differences in life expectancy decreases among non-Hispanic Black adults with insurance (0.1 years) and without insurance (0.1 years) (Table S15). Increases in years with uncontrolled hypertension were higher among non-Hispanic Black adults with insurance (0.2 years) compared to those without (0.1 years). However, differences by insurance status were smaller than in the scenario assuming racial heterogeneity in treatment effects.

## 4 Discussion

Our findings indicate that implementing antihypertensive treatment guidelines that no longer account for race may have adverse consequences for health outcomes and healthcare utilization among non-Hispanic Black adults, widening disparities with non-Hispanic white adults. However, the magnitude of these effects depended on assumptions of treatment heterogeneity by race.

Under assumptions of racial heterogeneity in treatment effectiveness, guidelines without race adjustment increased utilization of ACE inhibitors among non-Hispanic Black adults, a treatment class associated with smaller blood pressure reductions and higher discontinuation rates in this racial/ethnic group. This prescribing shift resulted in an average 0.4-year decrease in life expectancy and 0.8-year increase in years with uncontrolled hypertension among non-Hispanic Black adults, widening disparities with non-Hispanic white adults.

When treatment effectiveness was assumed equal across racial/ethnic groups, this same prescribing shift produced more modest effects, as ACE inhibitors were associated with smaller reductions in blood pressure compared to thiazide diuretics. Consequently, life expectancy decreased by 0.1 years, and years with uncontrolled hypertension increased by 0.2 years for non-Hispanic Black adults, less than one-half and one-quarter, respectively, of the effect observed under heterogeneous treatment assumptions. These results suggest that the consequences of guidelines without race adjustment for non-Hispanic Black adults are the most severe if racial heterogeneity in treatment effectiveness exists.

The existence of antihypertensive treatment heterogeneity by race is unclear. Beyond evidence from ALLHAT, analyses from the Veterans Affairs system, All of Us Research program, and additional clinical trials have demonstrated racial and ethnic differences in hypertension control across antihypertensive medication classes.^7,40,36–40^ However, it has not been established whether these differences are attributable to race, given evidence that race may not be a reliable proxy for genetic variation.^12,41^ Instead, observed treatment heterogeneity may be driven by differences in health insurance coverage, health literacy, medication adherence, or lower trust in the healthcare system.^2,42,43^

As a secondary analysis, we examined how incorporating heterogeneity in census tract-level SDI and health insurance coverage modified our findings. While non-Hispanic Black adults had lower rates of insurance coverage and higher census tract-level SDI values, we did not observe meaningful differences in life expectancy or hypertension control by racial groups compared to our main analysis. However, the adverse effects of guidelines that no longer account for race were more modest for non-Hispanic Black adults without insurance who had lower treatment access and thus less exposure to the prescribing shift towards ACE inhibitors. We argue that changes to clinical guidelines cannot be evaluated in isolation from the social context in which they are implemented.

Previous studies have examined the implications of removing race from clinical decision-making on health equity, most often in the context of race-adjusted clinical algorithms. These studies demonstrate that simply eliminating race from clinical algorithms does not necessarily improve and may, in some cases, exacerbate racial and ethnic disparities across clinical domains, including in chronic kidney disease, lung cancer, and colorectal cancer detection.^18,44–46^ Although several professional organizations have acknowledged the need to remove race from clinical practice guidelines,^47–49^ to our knowledge, no studies have evaluated the resulting implications for clinical decision-making, health outcomes, and racial/ethnic disparities.

Our analysis has several limitations stemming from the need to simplify some elements of a complex policy problem. First, we focused on antihypertensive monotherapy with either thiazide diuretics or ACE inhibitors. Antihypertensive treatment guidelines are more complex, allowing for additional medication classes such as angiotensin receptor blockers, calcium channel blockers, and beta blockers. Furthermore, patients who do not achieve blood pressure control often initiate combination therapy with multiple therapeutic options. Second, we did not explicitly model the effect of antihypertensive treatment on cardiovascular events, including stroke, congestive heart failure, and coronary heart disease.^28^ Further, we did not model any benefits from ACE inhibitors in slowing kidney disease progression.^50,51^ In our analysis, we assumed that antihypertensive treatment lowered blood pressure, which reduced the risk of all-cause mortality. Third, in our secondary analysis, we focused on incorporating health insurance status and census tract-level SDI. Additional social factors that may influence hypertension incidence and control were not incorporated. Finally, we focused our analysis on a population of non-Hispanic Black and non-Hispanic white adults to examine the implications of removing race-specific treatment recommendations for Black adults. We did not include additional racial and ethnic groups who also face disparities in hypertension, such as Hispanic and non-Hispanic Asian adults.^2,5^

## 5 Conclusion

Our study examined the implications of removing race adjustment from antihypertensive treatment guidelines on health outcomes, healthcare utilization, and disparities. We found that guidelines without race adjustment may result in adverse consequences for non-Hispanic Black adults. The magnitude of these effects depended on assumptions about whether racial heterogeneity in antihypertensive treatment effectiveness exists, which was the basis for previously published race-adjusted guidelines. Resolving the uncertainty around antihypertensive treatment heterogeneity and better understanding its underlying drivers are important when considering modification of clinical practice guidelines that may inadvertently widen racial and ethnic disparities in hypertension.

## Acknowledgements

Data for this project were accessed using the Stanford Center for Population Health Sciences Data Core. The PHS Data Core is supported by a National Institutes of Health National Center for Advancing Translational Science Clinical and Translational Science Award (UL1TR003142) and from Internal Stanford funding. The content is solely the responsibility of the authors and does not necessarily represent the official views of the NIH. We wish to acknowledge and thank the ABFM PRIME Registry participating clinicians and the American Board of Family Medicine, without whom the American Family Cohort would not be possible.

## Ethics statement

This study was approved by the Stanford University Institutional Review Board.

## Data availability

The data underlying this article cannot be shared publicly to protect the privacy of the individuals in the American Board of Family Medicine PRIME primary care electronic health record registry. The code to reproduce our results is available at: https://github.com/StanfordHPDS/hypertension-guideline-microsim

## Author contributions

Marika Cusick (Conceptualization, Methodology, Formal analysis, Visualization, Writing – original draft, Writing – review & editing), Malcolm Barrett (Formal analysis, Software, Writing – review & editing), Fernando Alarid-Escudero (Methodology, Writing – review & editing), Jeremy Goldhaber-Fiebert (Conceptualization, Formal analysis, Methodology, Writing – review & editing), Sherri Rose (Conceptualization, Methodology, Supervision, Writing – original draft, Writing – review & editing, Funding acquisition)

