## Supplement for "Hypertension disease natural history and healthcare utilization when treatment guidelines do not account for race"

### 6.1 Microsimulation model parameters

*Table S1: Model parameter dependencies and data sources*

| Parameter | Dependencies | Data |
|---|---|---|
| Simulation cohort | Age, sex, race/ethnicity, systolic and diastolic blood pressure values | NHANES[1] |
| *Disease natural history parameters* | | |
| Systolic blood pressure values | Age, sex | PRIME[2,3] |
| Diastolic blood pressure values | Age, sex | PRIME[2,3] |
| Hypertension mortality risk (hazard ratio) | Systolic and diastolic blood pressure values | Hansen et al.[4] |
| Baseline mortality rates | Age, sex, race/ethnicity | 2021 U.S. life tables[5] |
| *Health system utilization parameters* | | |
| Transition to detected/treated | Age, sex, systolic and diastolic blood pressure values | PRIME[2,3] |
| Transition to discontinued treatment | Type of antihypertensive therapy | ALLHAT[6] |
| *Antihypertensive treatment parameters* | | |
| Antihypertensive monotherapy utilization | Race/ethnicity (when using treatment guidelines that vary by race/ethnicity) | Holt et al.[7] |
| Antihypertensive treatment blood pressure reductions | Race/ethnicity (if heterogeneous effects are assumed), type of antihypertensive treatment | ALLHAT[6] |

*NHANES: National Health and Nutrition Examination Survey; ALLHAT: Antihypertensive and Lipid-Lowering Treatment to Prevent Heart Attack Trial*

### 6.2 Multivariate copula

To define our simulation cohort of interest, we used NHANES data from the 2013-2014, 2015-2016, and 2017-2018 years to fit a Gaussian multivariate copula. We selected a cohort of NHANES participants who were 18 years or older, self-identified as Non-Hispanic Black or non-Hispanic white, and self-reported not currently being on antihypertensive medication (n=6,939 survey participants). Continuous demographic and clinical variables of interest included age, family income to poverty ratio, number of healthcare visits in the past year, and systolic and diastolic blood pressure values. Binary variables of interest included sex (male or female), race/ethnicity (non-Hispanic Black or non-Hispanic white), and indicators for having a routine place for healthcare, having health insurance coverage, living below the poverty threshold, having less than 12 years of education, residing in a rented housing unit, being employed, living in a crowded housing unit, and being a single parent. We sampled one million synthetic individuals from the fitted multivariate copula and compared marginal distributions and correlation matrices between NHANES and the synthetic cohort (*Table S2*, *Table S3*). A Mantel test was conducted with a resulting p-value of <0.01 for the association between NHANES and synthetic cohort variable correlation matrices. This supports the preservation of NHANES correlation in our synthetic cohort. The final simulation cohort consisted of 50,000 non-Hispanic Black and 50,000 non-Hispanic white individuals aged 35 to 45 years, sampled at random from the synthetic copula cohort. Demographic and clinical characteristics by race/ethnicity are provided in *Table S4*. Individuals aged 35 to 45 years were sampled to increase the number of eligible synthetic individuals near the model starting age. All individuals were initialized at age 40 for the simulation. For comparability between the main and secondary analyses, we relied on the same simulation cohort.

*Table S2: Demographic and clinical characteristics of NHANES and multivariate copula synthetic cohort*

| Variable | Distribution | NHANES (%) | Synthetic Cohort (%) |
|---|---|---|---|
| Female | Bernoulli (p = 0.51) | 51% | 51% |
| Non-Hispanic Black | Bernoulli (p = 0.14) | 14% | 14% |
| Has place for routine healthcare | Bernoulli (p = 0.82) | 82% | 82% |
| Has health insurance | Bernoulli (p = 0.86) | 86% | 86% |
| Below poverty threshold | Bernoulli (p = 0.12) | 12% | 12% |
| Less than 12 years of education | Bernoulli (p = 0.08) | 8% | 8% |
| Rented housing unit | Bernoulli (p = 0.31) | 31% | 31% |
| Unemployed | Bernoulli (p = 0.06) | 6% | 6% |
| Living in a crowded housing unit | Bernoulli (p = 0.04) | 4% | 4% |
| Single parent | Bernoulli (p = 0.10) | 10% | 10% |
| **Variable** | **Distribution** | **NHANES (Mean [SD])** | **Synthetic Cohort (Mean [SD])** |
| Age | Uniform (min =19, max =80) | 44.4 [16.6] | 49.5 [17.6] |
| Ratio of family income to poverty | Gamma (shape = 3, rate = 0.75) | 3.2 [1.6] | 3.5 [1.4] |
| Times receiving healthcare in the past year | Negative binomial ($\mu$ =3.65, shape=5) | 3.7 [4.2] | 3.7 [2.5] |
| Systolic blood pressure | Normal (mean = 120, SD = 16) | 120.1 [15.5] | 120.1 [15.5] |
| Diastolic blood pressure | Normal (mean = 71, SD = 11) | 70.9 [10.9] | 70.9 [10.9] |

*NHANES: National Health and Nutrition Examination Survey; SD: Standard deviation*

*Table S3: Demographic and clinical correlation matrices in NHANES and multivariate copula synthetic cohorts*

| Variable 1 | Variable 2 | NHANES | Synthetic Cohort |
|---|---|---|---|
| Age | Female | 0.01 | 0.01 |
| Age | Times receiving healthcare over past year | 0.11 | 0.10 |
| Age | Single parent | -0.19 | -0.16 |
| Age | Unemployed | 0.04 | 0.02 |
| Age | Living in rented housing unit | -0.31 | -0.27 |
| Age | Has less than 12 years of education | 0.00 | 0.00 |
| Age | Below poverty threshold | -0.12 | -0.11 |
| Age | Living in a crowded housing unit | -0.12 | -0.09 |
| Age | Systolic blood pressure | 0.36 | 0.35 |
| Age | Ratio of family income to poverty | 0.18 | 0.17 |
| Age | Has health insurance | 0.14 | 0.15 |
| Age | Has place for routine healthcare | 0.22 | 0.23 |
| Age | Non-Hispanic Black | -0.13 | -0.11 |
| Age | Diastolic blood pressure | 0.10 | 0.10 |
| Below poverty threshold | Single parent | 0.16 | 0.11 |
| Below poverty threshold | Age | -0.12 | -0.11 |
| Below poverty threshold | Has place for routine healthcare | -0.07 | -0.03 |
| Below poverty threshold | Ratio of family income to poverty | -0.58 | -0.42 |
| Below poverty threshold | Systolic blood pressure | -0.00 | -0.00 |
| Below poverty threshold | Has less than 12 years of education | 0.22 | 0.16 |
| Below poverty threshold | Unemployed | 0.33 | 0.29 |
| Below poverty threshold | Living in a crowded housing unit | 0.20 | 0.15 |
| Below poverty threshold | Times receiving healthcare over past year | 0.04 | 0.02 |
| Below poverty threshold | Living in rented housing unit | 0.25 | 0.21 |
| Diastolic blood pressure | Below poverty threshold | -0.03 | -0.02 |
| Diastolic blood pressure | Female | -0.14 | -0.11 |
| Diastolic blood pressure | Systolic blood pressure | 0.48 | 0.48 |
| Diastolic blood pressure | Has less than 12 years of education | -0.01 | -0.01 |
| Diastolic blood pressure | Living in rented housing unit | -0.04 | -0.03 |
| Diastolic blood pressure | Unemployed | 0.02 | 0.01 |

| | | | |
|---|---|---|---|
| Diastolic blood pressure | Living in a crowded housing unit | -0.02 | -0.01 |
| Diastolic blood pressure | Single parent | -0.04 | -0.02 |
| Diastolic blood pressure | Times receiving healthcare over past year | -0.04 | -0.04 |
| Female | Living in rented housing unit | -0.02 | -0.01 |
| Female | Age | 0.01 | 0.01 |
| Female | Non-Hispanic Black | 0.01 | 0.00 |
| Female | Unemployed | 0.02 | 0.01 |
| Female | Has health insurance | 0.07 | 0.04 |
| Female | Has place for routine healthcare | 0.17 | 0.12 |
| Female | Living in a crowded housing unit | 0.00 | -0.00 |
| Female | Times receiving healthcare over past year | 0.16 | 0.13 |
| Female | Has less than 12 years of education | -0.05 | -0.02 |
| Female | Single parent | 0.13 | 0.11 |
| Female | Diastolic blood pressure | -0.14 | -0.11 |
| Female | Systolic blood pressure | -0.16 | -0.13 |
| Female | Ratio of family income to poverty | -0.04 | -0.03 |
| Female | Below poverty threshold | 0.04 | 0.02 |
| Has health insurance | Systolic blood pressure | -0.01 | -0.01 |
| Has health insurance | Living in a crowded housing unit | -0.10 | -0.07 |
| Has health insurance | Unemployed | -0.05 | -0.02 |
| Has health insurance | Living in rented housing unit | -0.15 | -0.13 |
| Has health insurance | Has less than 12 years of education | -0.15 | -0.10 |
| Has health insurance | Below poverty threshold | -0.20 | -0.17 |
| Has health insurance | Diastolic blood pressure | -0.00 | -0.00 |
| Has health insurance | Ratio of family income to poverty | 0.29 | 0.16 |
| Has health insurance | Times receiving healthcare over past year | 0.16 | 0.15 |
| Has health insurance | Single parent | -0.06 | -0.03 |
| Has health insurance | Has place for routine healthcare | 0.25 | 0.24 |
| Has less than 12 years of education | Times receiving healthcare over past year | -0.00 | -0.00 |
| Has less than 12 years of education | Single parent | 0.07 | 0.02 |
| Has less than 12 years of education | Living in a crowded housing unit | 0.12 | 0.07 |

| | | | |
|---|---|---|---|
| Has less than 12 years of education | Living in rented housing unit | 0.08 | 0.04 |
| Has less than 12 years of education | Below poverty threshold | 0.22 | 0.16 |
| Has less than 12 years of education | Ratio of family income to poverty | -0.26 | -0.22 |
| Has less than 12 years of education | Unemployed | 0.13 | 0.08 |
| Has less than 12 years of education | Has health insurance | -0.15 | -0.10 |
| Has less than 12 years of education | Female | -0.05 | -0.02 |
| Has place for routine healthcare | Systolic blood pressure | 0.00 | 0.00 |
| Has place for routine healthcare | Female | 0.17 | 0.12 |
| Has place for routine healthcare | Non-Hispanic Black | -0.05 | -0.02 |
| Has place for routine healthcare | Has health insurance | 0.25 | 0.24 |
| Has place for routine healthcare | Ratio of family income to poverty | 0.14 | 0.09 |
| Has place for routine healthcare | Diastolic blood pressure | -0.01 | -0.01 |
| Has place for routine healthcare | Below poverty threshold | -0.07 | -0.03 |
| Has place for routine healthcare | Has less than 12 years of education | -0.09 | -0.08 |
| Has place for routine healthcare | Living in rented housing unit | -0.12 | -0.12 |
| Has place for routine healthcare | Unemployed | -0.01 | -0.00 |
| Has place for routine healthcare | Living in a crowded housing unit | -0.05 | -0.02 |
| Has place for routine healthcare | Single parent | -0.02 | -0.01 |
| Has place for routine healthcare | Times receiving healthcare over past year | 0.21 | 0.26 |
| Living in a crowded housing unit | Single parent | 0.14 | 0.10 |
| Living in a crowded housing unit | Living in rented housing unit | 0.15 | 0.11 |
| Living in a crowded housing unit | Below poverty threshold | 0.20 | 0.15 |
| Living in a crowded housing unit | Times receiving healthcare over past year | -0.01 | -0.00 |
| Living in rented housing unit | Times receiving healthcare over past year | -0.03 | -0.02 |
| Living in rented housing unit | Single parent | 0.12 | 0.11 |
| Living in rented housing unit | Age | -0.31 | -0.27 |
| Living in rented housing unit | Unemployed | 0.07 | 0.03 |
| Living in rented housing unit | Has place for routine healthcare | -0.12 | -0.12 |
| Living in rented housing unit | Non-Hispanic Black | 0.24 | 0.21 |
| Living in rented housing unit | Female | -0.02 | -0.01 |
| Living in rented housing unit | Living in a crowded housing unit | 0.15 | 0.11 |
| Non-Hispanic Black | Age | -0.13 | -0.11 |

| | | | |
|---|---|---|---|
| Non-Hispanic Black | Has place for routine healthcare | -0.05 | -0.02 |
| Non-Hispanic Black | Has health insurance | -0.14 | -0.09 |
| Non-Hispanic Black | Ratio of family income to poverty | -0.23 | -0.19 |
| Non-Hispanic Black | Systolic blood pressure | 0.06 | 0.04 |
| Non-Hispanic Black | Has less than 12 years of education | 0.09 | 0.08 |
| Non-Hispanic Black | Below poverty threshold | 0.19 | 0.16 |
| Non-Hispanic Black | Living in rented housing unit | 0.24 | 0.21 |
| Non-Hispanic Black | Unemployed | 0.06 | 0.02 |
| Non-Hispanic Black | Living in a crowded housing unit | 0.13 | 0.10 |
| Non-Hispanic Black | Times receiving healthcare over past year | -0.08 | -0.05 |
| Non-Hispanic Black | Diastolic blood pressure | 0.01 | 0.01 |
| Non-Hispanic Black | Single parent | 0.20 | 0.16 |
| Ratio of family income to poverty | Living in rented housing unit | -0.36 | -0.36 |
| Ratio of family income to poverty | Unemployed | -0.28 | -0.21 |
| Ratio of family income to poverty | Below poverty threshold | -0.58 | -0.42 |
| Ratio of family income to poverty | Has less than 12 years of education | -0.26 | -0.22 |
| Ratio of family income to poverty | Diastolic blood pressure | 0.06 | 0.05 |
| Ratio of family income to poverty | Living in a crowded housing unit | -0.22 | -0.15 |
| Ratio of family income to poverty | Single parent | -0.20 | -0.18 |
| Ratio of family income to poverty | Times receiving healthcare over past year | 0.01 | 0.01 |
| Ratio of family income to poverty | Systolic blood pressure | -0.02 | -0.02 |
| Single parent | Age | -0.19 | -0.16 |
| Single parent | Has health insurance | -0.06 | -0.03 |
| Single parent | Ratio of family income to poverty | -0.20 | -0.18 |
| Single parent | Times receiving healthcare over past year | -0.02 | -0.01 |
| Single parent | Living in rented housing unit | 0.12 | 0.11 |
| Single parent | Below poverty threshold | 0.16 | 0.11 |
| Systolic blood pressure | Age | 0.36 | 0.35 |
| Systolic blood pressure | Living in a crowded housing unit | -0.03 | -0.01 |
| Systolic blood pressure | Unemployed | 0.03 | 0.02 |
| Systolic blood pressure | Times receiving healthcare over past year | -0.03 | -0.03 |
| Systolic blood pressure | Living in rented housing unit | -0.06 | -0.05 |
| Systolic blood pressure | Has less than 12 years of education | 0.05 | 0.03 |

| | | | |
|---|---|---|---|
| Systolic blood pressure | Below poverty threshold | -0.00 | -0.00 |
| Systolic blood pressure | Diastolic blood pressure | 0.48 | 0.48 |
| Systolic blood pressure | Ratio of family income to poverty | -0.02 | -0.02 |
| Systolic blood pressure | Single parent | -0.08 | -0.05 |
| Times receiving healthcare over past year | Ratio of family income to poverty | 0.01 | 0.01 |
| Times receiving healthcare over past year | Single parent | -0.02 | -0.01 |
| Times receiving healthcare over past year | Has less than 12 years of education | -0.00 | -0.00 |
| Unemployed | Single parent | 0.04 | 0.01 |
| Unemployed | Living in a crowded housing unit | 0.04 | 0.01 |
| Unemployed | Living in rented housing unit | 0.07 | 0.03 |
| Unemployed | Systolic blood pressure | 0.03 | 0.02 |
| Unemployed | Has health insurance | -0.05 | -0.02 |
| Unemployed | Times receiving healthcare over past year | 0.06 | 0.03 |
| Unemployed | Has less than 12 years of education | 0.13 | 0.08 |

*NHANES: National Health and Nutrition Examination Survey*

*Table S4: Simulation cohort characteristics by racial group*

| Variable | Non-Hispanic Black (%) | Non-Hispanic white (%) |
|---|---|---|
| Female | 52% | 51% |
| Has place for routine healthcare | 79% | 79% |
| Has health insurance | 79% | 86% |
| Below poverty threshold | 26% | 11% |
| Less than 12 years of education | 14% | 7% |
| Rented housing unit | 57% | 31% |
| Unemployed | 7% | 6% |
| Living in a crowded housing unit | 8% | 4% |
| Single parent | 22% | 9% |
| **Variable** | **Non-Hispanic Black (Mean [SD])** | **Non-Hispanic white (Mean [SD])** |
| Age | 39.9 [2.9] | 40.0 [2.9] |
| Ratio of family income to poverty | 2.7 [1.4] | 3.5 [1.4] |
| Times receiving healthcare over the past year | 3.3 [2.4] | 3.6 [2.5] |
| Systolic blood pressure | 120.9 [14.4] | 117.2 [14.5] |
| Diastolic blood pressure | 71.0 [11.0] | 70.4 [10.8] |

*SD: standard deviation*

### 6.3 Systolic and diastolic quantile regression estimation

We estimated systolic and diastolic blood pressure trajectories using the American Board of Family Medicine PRIME registry. From the PRIME registry data, we identified all systolic and diastolic blood pressure measurements between January 2018 and June 2024 prior to any antihypertensive medication exposure (23 million measurements for 3.6 million patients). The following first-line antihypertensive drugs were used to identify exposure: chlorthalidone, hydrochlorothiazide, indapamide, metolazone, benazepril, captopril, fosinopril, lisinopril, moexipril, perindopril, ramipril, quinapril, trandolapril, azilsartan, candesartan, eprosartan, irbesartan, losartan, olmesartan, telmisartan, valsartan, amlodipine, felodipine, isradipine, nicardipine, nifedipine, nisoldipine, diltiazem, and verapamil.[8]

A subset of one million blood pressure observations (n=695,714 patients) was randomly selected to fit quantile regressions for predicting systolic and diastolic blood pressure values. Quantile regressions ranged between the 1st and 99th quantiles, as the 0th and 100th quantiles were not supported by the Python statsmodels package. In the simulation, each individual was assigned systolic and diastolic quantiles, and the corresponding quantile regression model was used to generate that individual's predicted blood pressure value at each simulated age.

Systolic blood pressure $Y_{it}$ was estimated as a function of age $(X_{it})$, sex $(F_i)$, and an interaction between patient age and sex $(F_i X_{it})$ for an individual $i$ at time $t$:

$$Y_{it} = \beta_0 + \sum_{j=1}^{k=6} \beta_j\, B(X_{it}) + \beta_6 F_i + \sum_{j=5}^{k=11} \beta_j\, F_i B(X_{it}).$$

Age was modeled using a linear B-spline basis with 5 degrees of freedom. Spline specification and degrees of freedom were selected via 5-fold cross-validation to minimize quantile loss.

We estimated quantile regressions to predict diastolic blood pressure values $Z_{it}$ as a function of age, sex, and an interaction between patient age and sex:

$$Z_{it} = \beta_0 + \sum_{j=1}^{k=6} \beta_j\, B(X_{it}) + \beta_7 F_i + \sum_{j=8}^{k=13} \beta_j\, F_i B(X_{it}).$$

Age was modeled using a cubic B-spline basis with 6 degrees of freedom, selected via 5-fold cross-validation.

In our secondary analysis, we incorporated census tract-level social deprivation index (SDI) $(S_i)$ in predicting systolic blood pressure values:

$$Y_{it} = \beta_0 + \sum_{j=1}^{k=5} \beta_j\, B(X_{it}) + \beta_6 F_i + \beta_7 S_i + \sum_{j=8}^{k=12} \beta_j\, F_i B(X_{it}),$$

and diastolic blood pressure values:

$$Z_{it} = \beta_0 + \sum_{j=1}^{k=6} \beta_j\, B(X_{it}) + \beta_7 F_i + \beta_8 S_i + \sum_{j=9}^{k=14} \beta_j\, F_i B(X_{it}).$$

Selected using 5-fold cross-validation, we modeled age using a quadratic B-spline basis with 5 degrees of freedom and a cubic B-spline basis with 6 degrees of freedom for our systolic and diastolic quantile regressions, respectively. The average R-squared values for the systolic and diastolic blood pressure regressions were 0.11 and 0.08, respectively.[9]

Our quantile regression approach has several limitations. First, we estimated the quantile regressions using PRIME registry data with repeated measurements from individuals, assuming independence between observations. Ignoring dependence between repeated measurements can impact standard error estimates of our regression model predictions. However, our simulation analysis does not rely on these standard error estimates. An alternative approach could consider the use of hierarchical quantile regression models. Second, while census tract-level SDI can vary over time given individual mobility and neighborhood changes, we only had one census tract-level SDI value per individual in the PRIME registry data. Thus, census tract-level SDI is time-invariant in the quantile regressions pertaining to our secondary analysis.

### 6.4 Systolic and diastolic blood pressure quantile sampling

To simulate individual blood pressure trajectories, we assumed that individual systolic
To simulate individual blood pressure trajectories, we assumed that individual systolic
and diastolic blood pressure quantiles varied over time. These sampled quantiles determined which quantile regression model was used to predict a simulated individual's monthly systolic and diastolic blood pressure values. To allow for individual variation in quantile switching, we updated person-specific quantiles using a stochastic process with a mean interval of five years.

Correlations between updated quantiles were estimated using PRIME data. Using all systolic and diastolic blood pressure measurements prior to any antihypertensive medication exposure in the PRIME registry, we ordered blood pressure measurements by measurement date and identified the previously recorded blood pressure value for each patient. To focus on regularly spaced measurements, we excluded all blood pressure measurements for which the previous measurement occurred more than six months earlier (n=1.9 million patients). From this dataset, we randomly sampled one million observations (n=205,608 patients) to estimate blood pressure quantile correlations.

We estimated correlations using the following procedure. First, for each blood pressure observation, we calculated the number of years since the patient's first recorded measurement ($t = 0$). Second, for each observation, we computed the systolic and diastolic blood pressure percentiles relative to observations from patients of the same age and sex. Third, we transformed the data such that each patient had a single row, with columns representing their average age-sex-specific blood pressure percentiles for each observed year. For example, a patient $i$ with four years of blood pressure measurements would have eight percentiles: one systolic ($SP_{i,t}$) and diastolic percentile ($DP_{i,t}$) for each year $t$. Fourth, we estimated correlation matrices for patient-specific percentiles across 11 age groups, divided into four-year intervals (e.g., 40–43 years, 44–47 years). Finally, to assign percentile changes over time, we assumed that correlations from different age cohorts replicated those of a single cohort observed over time. In *Table S5*, we reported observed correlations between systolic percentiles at year one and systolic and diastolic percentiles in the following three years.

We assigned each individual a sequence of systolic and diastolic quantile update months by randomly sampling quantile switching intervals from an exponential distribution with a mean of five years. This resulted in individual-specific quantile update schedules over the lifetime horizon. In our secondary analysis, we followed the same procedure, but computed blood pressure percentile correlations based on age, sex, and census tract-level SDI.

*Table S5: Correlations between age-sex-specific systolic and diastolic percentiles during four years of observation in PRIME registry*

| Correlation | | Value |
|---|---|---|
| Systolic blood pressure (year 1) | Systolic blood pressure (year 2) | 0.57 |
| Systolic blood pressure (year 1) | Systolic blood pressure (year 3) | 0.49 |
| Systolic blood pressure (year 1) | Systolic blood pressure (year 4) | 0.48 |
| Systolic blood pressure (year 1) | Diastolic blood pressure (year 1) | 0.68 |
| Systolic blood pressure (year 1) | Diastolic blood pressure (year 2) | 0.46 |
| Systolic blood pressure (year 1) | Diastolic blood pressure (year 3) | 0.40 |
| Systolic blood pressure (year 1) | Diastolic blood pressure (year 4) | 0.36 |

### 6.5 Mortality adjustments by systolic and diastolic blood pressure values

Baseline mortality rates were obtained from age-sex-racial/ethnic-group-specific 2021 U.S. life tables.[5] We adjusted baseline mortality rates by systolic and diastolic blood pressure values using previously published all-cause mortality hazard ratio estimates (*Table S6*).[4] We assumed that hazard ratios remained constant outside the ranges estimated in this study: above 90 mm Hg for diastolic blood pressure and above 160 mm Hg for systolic blood pressure. Because the published estimates reported separate univariate hazard ratios for systolic and diastolic blood pressure, we used their average as an approximation of blood pressure-related mortality risk.[4]

To account for the high prevalence of hypertension in the U.S. population, we estimated age-group-specific mean systolic and diastolic blood pressures in five-year intervals (e.g., 40-to-44 year-olds, 45-to-49 year-olds, etc.) using NHANES data among U.S. adults who self-reported as non-Hispanic Black or non-Hispanic white. We then linearly interpolated these estimates to obtain age-specific mean systolic and diastolic blood pressure values for each simulated age. We assumed constant mean systolic and diastolic blood pressure values for individuals aged 80 years and older, given limited sample sizes of older individuals in NHANES. Using these average systolic and diastolic values, we calculated the average blood-pressure-related hazard increase at each age. For each simulated individual, we scaled their blood-pressure-related hazard increase by dividing it by the age-specific average blood pressure hazard increase. This scaling ensured that the average mortality rate at each age remained anchored to the U.S. life table. Individuals with higher or lower blood pressure than the age-specific average experienced correspondingly higher or lower mortality risk.

*Table S6: Univariate hazard ratios for all-cause mortality risk by systolic and diastolic blood pressure values (mm Hg)*[4]

| Systolic Blood Pressure | Hazard Ratio | Diastolic Blood Pressure | Hazard Ratio |
|---|---|---|---|
| 115 | 1.0 | 65 | 1.0 |
| 124 | 2.0 | 72 | 1.8 |
| 130 | 2.0 | 76 | 2.0 |
| 140 | 2.4 | 82 | 2.0 |
| 155 | 4.0 | 90 | 3.0 |

### 6.6 Receiving antihypertensive treatment

Treatment initiation was modeled as a two-step process: simulated individuals first had to have a blood pressure measurement, and then, during that visit, could initiate antihypertensive therapy. Our model assumed that only patients who were in the health system with blood pressure values corresponding to at least stage 1 hypertension (systolic blood pressure: ≥130 mm Hg or diastolic blood pressure: ≥80 mm Hg) were eligible to receive antihypertensive treatment. We predicted the number of months until the next measurement visit ($M_{i,t+1}$) using quantile regressions as a function of age, sex, systolic blood pressure, and diastolic blood pressure:

$$M_{i,t+1} = \beta_0 + \beta_1 X_{it} + \beta_2 F_i + \beta_3 Y_{it} + \beta_4 Z_{it}.$$

We fit our quantile regressions to PRIME registry data, using a similar approach described in *Section 6.4*. One million observations from 662,095 patients were randomly sampled, including measures that occurred more than six months apart. The number of months between blood pressure measurements was on average 5.5 months [6.0]. The average pseudo R-squared value of our quantile regression was less than 0.01.

We assigned each simulated individual a measurement frequency quantile to predict the number of months until their next measurement using NHANES data. Because NHANES does not provide longitudinal blood pressure measurement frequencies, self-reported healthcare visits were used to assign individuals to relative measurement-frequency quantiles. We used the NHANES survey question, “How many times have you seen a doctor or other healthcare professional about your health at a doctor’s office, a clinic or some other place? Do not include times you were hospitalized overnight, visits to hospital emergency rooms, home visits or telephone calls.” In our NHANES cohort, the average number of self-reported healthcare visits was 3.4 [2.4] per year. We assumed that blood pressure measurements occurred during these visits and transformed visit frequencies into months between measurements. After transformation, the average number of assigned months between measurements was 4.7 months [4.6].

For each simulated individual, we determined the measurement frequency quantile regression that most closely approximated their assigned number of months since their previous measurement. This quantile was then used to predict the simulated frequency of measurement visits over their lifetime. We compared the distribution of months between measurements in our simulated cohort and a random subset of 10,000 observations corresponding to 40-year-old adults in the PRIME registry (*Figure S1*).

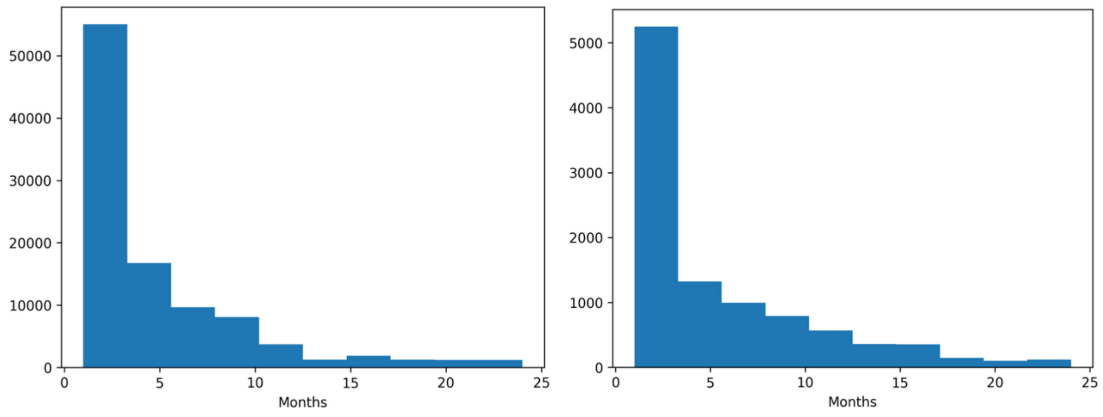


*(a)* *(b)*

*Figure S1: Number of months between blood pressure measurements for 40-year-olds in (a) model simulation cohort and (b) PRIME registry*

At a measurement visit, simulated individuals with a blood pressure value corresponding to at least stage 1 hypertension may receive antihypertensive treatment. We predicted the probability of initiating antihypertensive treatment at a given measurement visit $P(A_{it} = 1)$ using a logistic regression as a function of patient age, sex, and systolic and diastolic blood pressure values:

$$\text{logit}(P(A_{it} = 1)) = \beta_0 + \beta_1 X_{it} + \beta_2 F_i + \beta_3 Y_{it} + \beta_4 Z_{it}.$$

Our logistic regression was estimated using PRIME registry blood pressure observations with either systolic blood pressure of ≥130 mm Hg or diastolic blood pressure of ≥80 mm Hg prior to antihypertensive treatment. We identified a total of 10.7 million observations for 2.4 million patients and randomly sampled one million observations (n=404,908 patients) to fit the logistic regression. If a simulated individual did not initiate treatment at a measurement visit, we predicted the number of months until their next measurement and repeated this procedure until the simulated individual initiated treatment or died.

To validate rates of antihypertensive treatment exposure in our model, we estimated a Kaplan-Meier curve of individuals in the PRIME registry receiving antihypertensive treatment over time. Because we only had up to 6 years of follow-up in our data, we constructed a longer-term Kaplan-Meier curve using left-truncated observations. This approach allowed individuals entering observation at older ages to contribute person-time only after their entry age, thereby approximating a longer age-based treatment initiation curve.

Assuming an index date of January 1, 2018, we followed cohorts of non-Hispanic Black and non-Hispanic white individuals aged 40, 46, 52, 58, 64, or 70 years who had not been exposed to antihypertensive treatment at the index date. We assumed the age of entry was the age of the age-specific cohort (e.g., 40 for the 40-year-old cohort). Exit age was defined as the minimum of the age at antihypertensive treatment exposure, age at last observed blood pressure measurement, and age at death simulated from age-sex-race-specific U.S. life tables. Observations were reweighted so that 50% of the adults were non-Hispanic Black and 50% were non-Hispanic white, matching the composition of our simulation cohort. The cumulative density of the Kaplan-Meier curve was compared with the fraction of alive adults in the detected and treated health system or discontinued health system states in our simulation model. Our model underestimated the cumulative incidence of antihypertensive treatment exposure observed in the PRIME registry. The high observed cumulative incidence in PRIME likely reflects individuals who initiated antihypertensive treatment for other health benefits beyond hypertension control, whereas our simulation model assumed that patients may initiate treatment only with elevated blood pressure values (*Figure S2*).

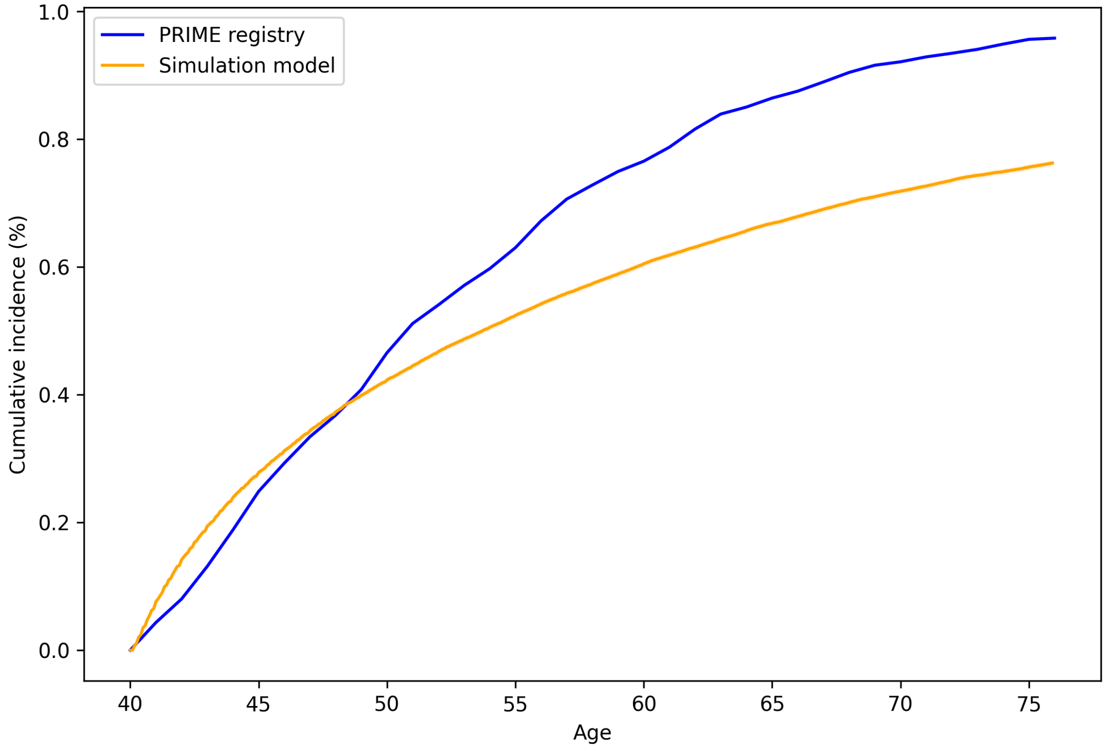


*Figure S2: Comparing the cumulative incidence of antihypertensive treatment exposure between PRIME registry and simulation cohort*

We also compared the cumulative incidence of antihypertensive treatment in our model with published estimates from the Framingham Heart Study.[10] The study reported the cumulative probability of receiving antihypertensive therapy from age 55 at 10, 15, 20, and 25 years of follow-up. As demonstrated in *Figure S3*, our simulation model's cumulative incidence aligned closely with the published estimates at 15 and 20 years. However, the simulation model overestimated the cumulative incidence of treatment exposure at 10 years and underestimated it at 25 years.

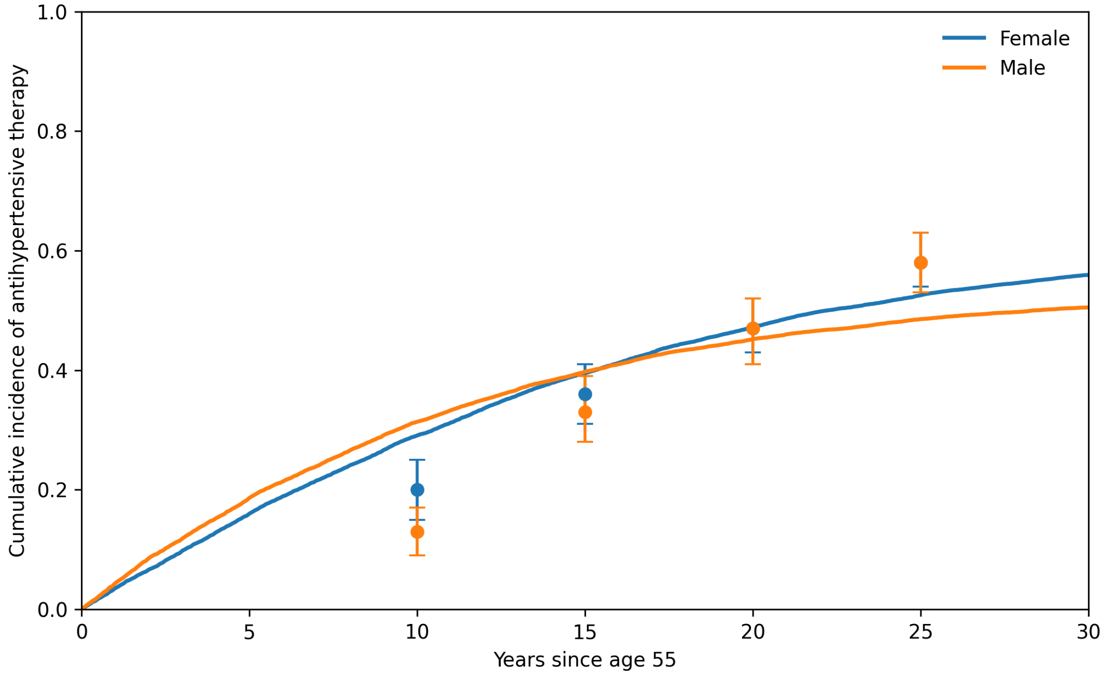


*Figure S3: Comparing the cumulative incidence of antihypertensive treatment exposure in the Framingham Heart Study and simulation cohort*

In our secondary analysis, we incorporated census tract-level SDI in predicting the number of months until the next measurement visit:

$$M_{i,t+1} = \beta_0 + \beta_1 X_{it} + \beta_2 F_i + \beta_3 S_i + \beta_4 Y_{it} + \beta_5 Z_{it},$$

and the probability of initiating antihypertensive treatment:

$$\text{logit}(P(A_{it} = 1)) = \beta_0 + \beta_1 X_{it} + \beta_2 F_i + \beta_3 S_i + \beta_4 Y_{it} + \beta_5 Z_{it}.$$

### 6.7 Validation analysis results

We validated the baseline simulation model against observed age-specific hypertension stage prevalence among individuals in the PRIME registry prior to antihypertensive medication use. In the baseline version of the model, we did not model health system utilization and thus did not simulate the effect of treatment on blood pressure. We sampled a validation cohort of 10,000 40-year-old patients from the PRIME registry. Demographic and clinical characteristics of our validation cohort are in *Table S7*.

*Table S7: Demographic and clinical characteristics of PRIME registry validation cohort*

| Categorical Variables | | Category (%) |
|---|---|---|
| Race/ethnicity | Hispanic | 12% |
| | Non-Hispanic Black | 5% |
| | Non-Hispanic white | 49% |
| | Other race/ethnicity | 34% |
| Sex | Female | 63% |
| Initial hypertension stage | Normal | 34% |
| | Elevated | 15% |
| | Stage 1 | 35% |
| | Stage 2 | 16% |
| **Continuous Variables** | | **Mean [SD]** |
| Census tract-level social deprivation index | | 14.6 [8.2] |
| Systolic blood pressure | | 122.1 [13.3] |
| Diastolic blood pressure | | 77.4 [9.3] |

*SD: standard deviation*

The average life expectancy of our 40-year-old cohort was 38.2 years (95% CI: 38.0, 38.5). In *Table S8*, we observed concordance between the average life expectancies computed by our simulation model and life expectancies by race and ethnicity reported in the 2021 U.S. life tables.[5]

*Table S8: Comparison of mean life expectancies in validation cohort with U.S. life table estimates by race/ethnicity*

| Race/Ethnicity | Validation Cohort, Mean [MCSE] | 2021 U.S. Life Table |
|---|---|---|
| Hispanic | 39.4 [<0.01] | 40.1 |
| Non-Hispanic Black | 35.3 [<0.01] | 35.3 |
| Non-Hispanic white | 38.6 [<0.01] | 38.9 |

*MCSE: Monte Carlo standard error*

*Figure S4* shows concordance between age-specific hypertension stage prevalence in the simulation cohort and the observed PRIME registry estimates, with 95% confidence intervals.

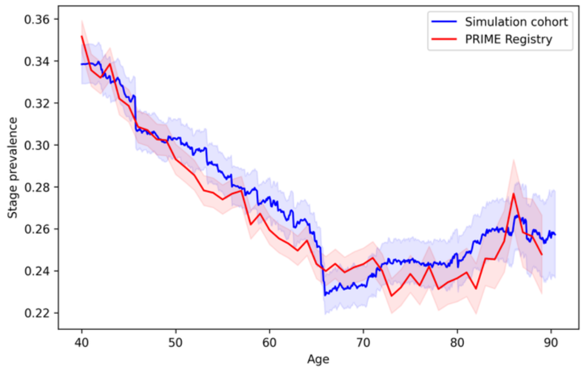


(a)

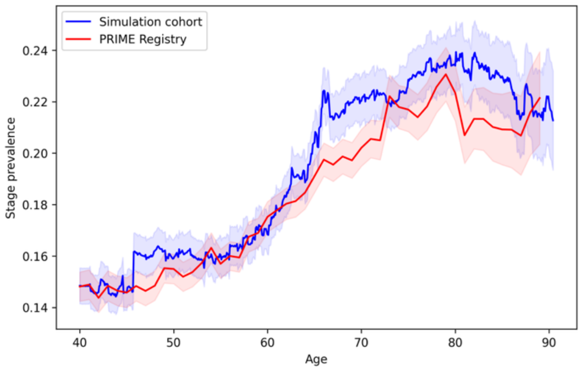


(b)

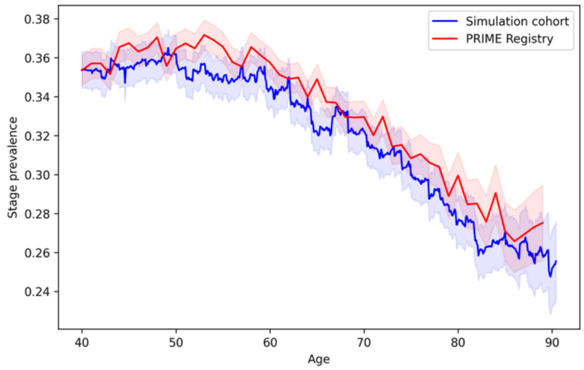


*(c)*

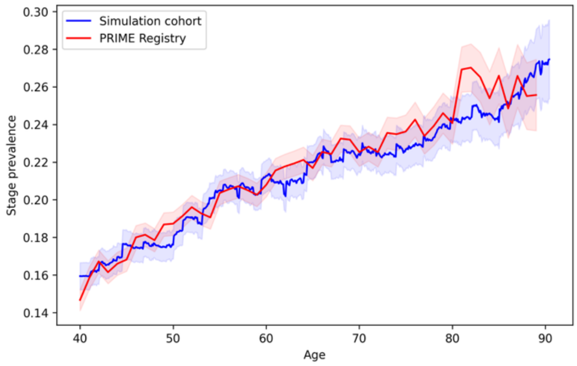


*(d)*

*Figure S4: Comparing the age-specific hypertension stage prevalence between the PRIME registry and simulation cohort: (a) Normal, (b) Elevated, (c) Stage 1, and (d) Stage 2*

We also compared the lifetime incidence of hypertension from our simulation model with a previously published study, which estimated the lifetime risk of hypertension for Black women, Black men, white women, and white men.[11] Estimates and 95% confidence intervals from the published study are plotted with dots and vertical error bars. Our simulation model overestimated hypertension incidence among all groups, particularly among white women.

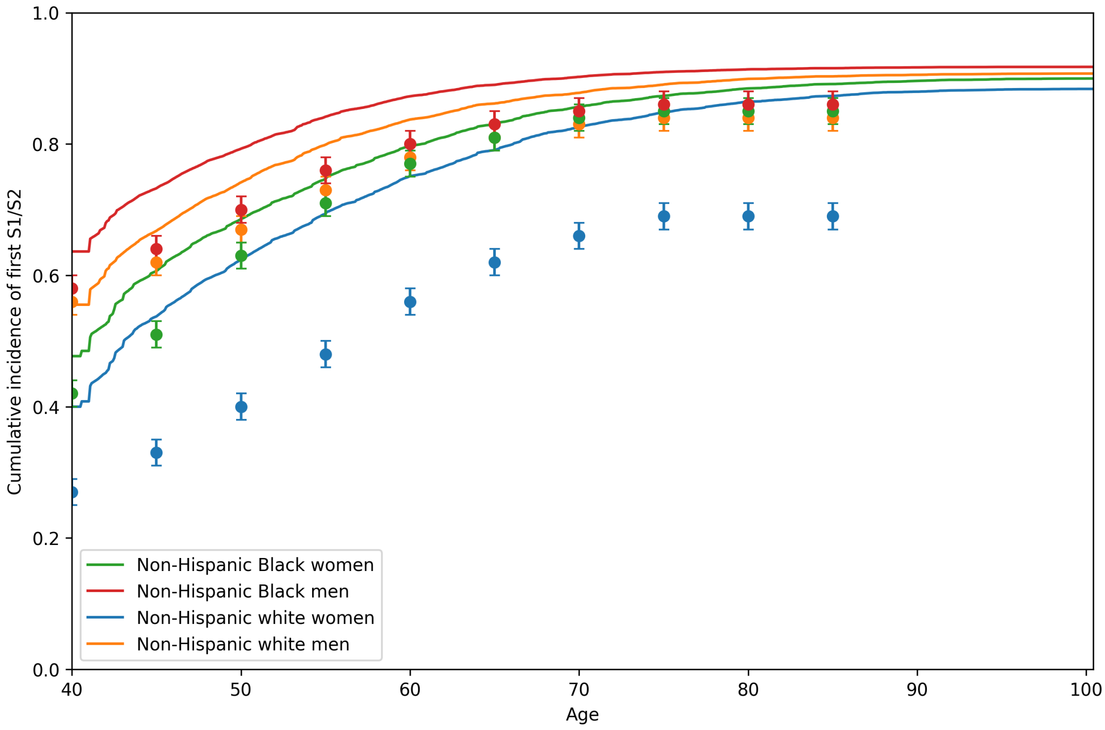


*Figure S5: Cumulative incidence of hypertension (defined as systolic blood pressure ≥ 130 mm Hg or diastolic blood pressure ≥ 80 mm Hg) from our simulation model (lines) and published study (dots)*

### 6.8 Secondary analysis with the social factors framework

Using a previously developed social factors framework,[12] we modeled the impact of two social factors: health insurance status and census tract-level SDI, as demonstrated in the model diagram (*Figure S6*) and model transition probabilities (*Table S9*). Sections *Section 6.9*, *Section 6.10*, *Section 6.11*, and *Section 6.12* describe additional changes to the model for our secondary analysis with the social factors framework.

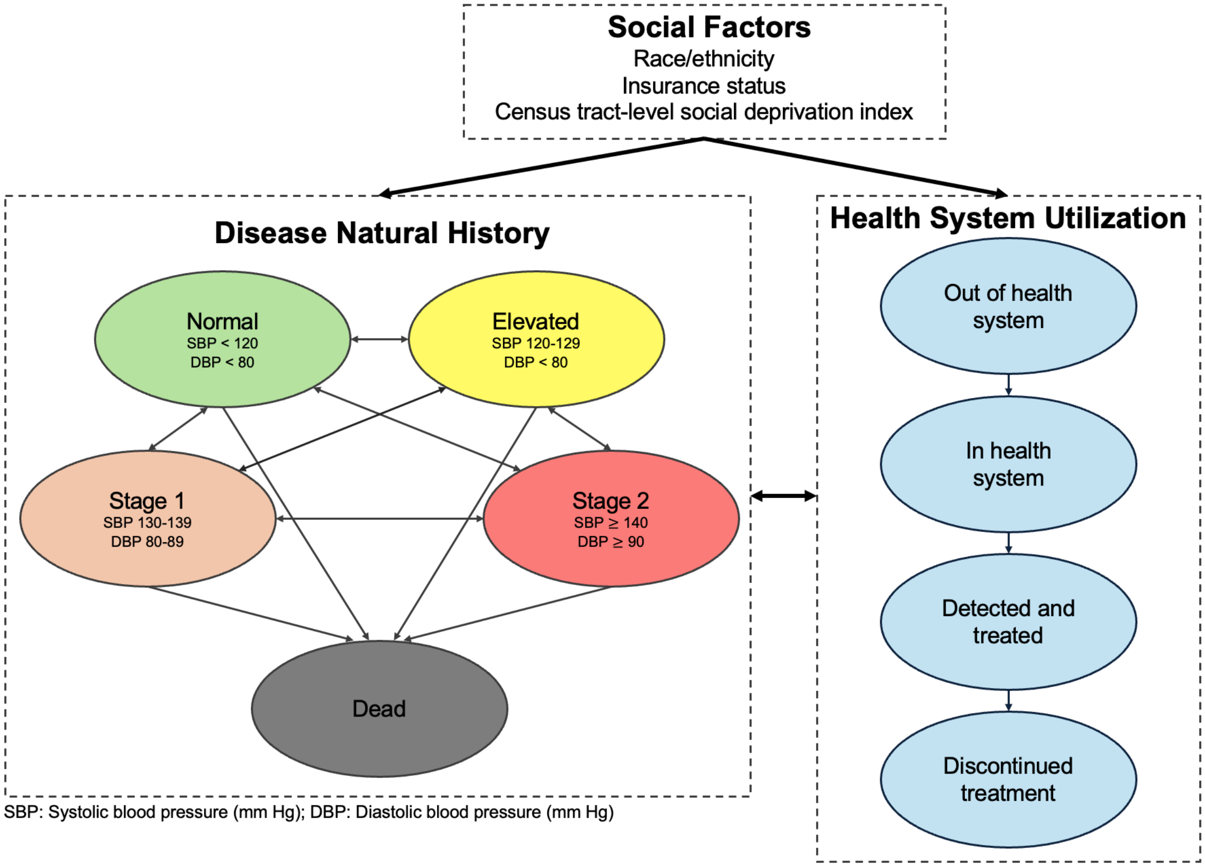


*Figure S6: Model diagram of state-transition microsimulation model with social factors framework*

*Table S9: Model parameter dependencies and data sources for secondary analysis*

| Parameter | Dependencies | Data |
|---|---|---|
| Simulation cohort | Age, sex, race/ethnicity, place for routine healthcare, insurance status, systolic and diastolic blood pressure values | NHANES[1] |
| *Disease natural history parameters* | | |
| Systolic blood pressure values | Age, sex, SDI | PRIME[2,3] |
| Diastolic blood pressure values | Age, sex, SDI | PRIME[2,3] |
| Systolic and diastolic blood pressure increases | Health system status (out of health system) | NHANES[1] |
| Hypertension mortality risk (hazard ratio) | Systolic and diastolic blood pressure values | Hansen et al.[4] |
| Baseline mortality rates | Age, sex, race/ethnicity | 2021 U.S. life tables[5] |
| Increased mortality risk from being uninsured (hazard ratio) | | Wilper et al.[13] |
| *Health system utilization parameters* | | |
| Transition to in health system | Insurance status | NHANES[1] |
| Transition to detected/treated | Age, sex, SDI, systolic and diastolic blood pressure values | PRIME[2,3] |
| Transition to discontinued treatment | Insurance status, type of antihypertensive therapy | ALLHAT[6] |
| *Antihypertensive treatment parameters* | | |
| Antihypertensive monotherapy utilization | Race/ethnicity (when using treatment guidelines that vary by race/ethnicity) | Holt et al.[7] |
| Antihypertensive treatment blood pressure reductions | Race/ethnicity (if heterogeneous effects are assumed), type of antihypertensive treatment | ALLHAT[6] |

*NHANES: National Health and Nutrition Examination Survey; ALLHAT: Antihypertensive and Lipid-Lowering Treatment to Prevent Heart Attack Trial*

### 6.9 Census tract-level social deprivation index assignment

Our simulation analysis required census tract-level SDI to predict model transition probabilities. However, the publicly available NHANES data used to create our simulation cohort did not include geocoded census tract information. To assign census tract-level SDI values to individuals in our cohort, we applied a Bayesian approach based on individual-level characteristics from NHANES. Individuals with a characteristic (e.g., being unemployed) were more likely to be assigned to a census tract with a higher percentage of individuals unemployed and younger than 65 years of age. The SDI metric comprises several area-level measures, including the percentages of individuals living in poverty, having less than 12 years of education, living in a single-parent household, residing in rented housing units, living in overcrowded housing units, lacking access to a car, and being unemployed and younger than 65 years of age.[14] NHANES provided individual-level data for all SDI components with the exception of car ownership.

We obtained 2023 SDI data on 83,612 census tracts, which included population counts and the estimated fraction of individuals with each SDI component in the census tract. Using census tract population prevalence as the prior probability, we calculated each individual's posterior probability of belonging to each census tract based on SDI-related characteristics observed in NHANES. Using these posterior probabilities, we sampled ten census tracts and assigned the average SDI value from the sampled census tracts as the individual's census tract-level SDI value. Averaging across multiple census tracts reduced the influence of any single extreme census tract-level SDI value.

In our simulation cohort, assigned census tract-level SDI values were on average 16.3 with a standard deviation of 6.0. Among those with insurance coverage, average assigned census tract-level SDI values were lower (15.9 [5.7]) compared to those without insurance coverage (18.4 [6.7]).

To ensure comparability with data from the PRIME registry, we validated our assigned census tract-level SDI values by comparing them to census tract-level SDI values of a randomly selected subset of 10,000 40-year-old individuals from the PRIME registry (5,000 non-Hispanic Black and 5,000 non-Hispanic white patients). Average census tract-level SDI was 16.1 with a standard deviation of 8.9 in the PRIME registry. *Figure S7* shows histograms of census tract-level SDI values for our main simulation cohort and the randomly sampled subset from the PRIME registry. While census tract-level SDI values can vary over time, we assumed a single census tract-level SDI value per individual over their lifetime. The PRIME registry only has a single census tract-level SDI per individual and does not include historical census-tract data.

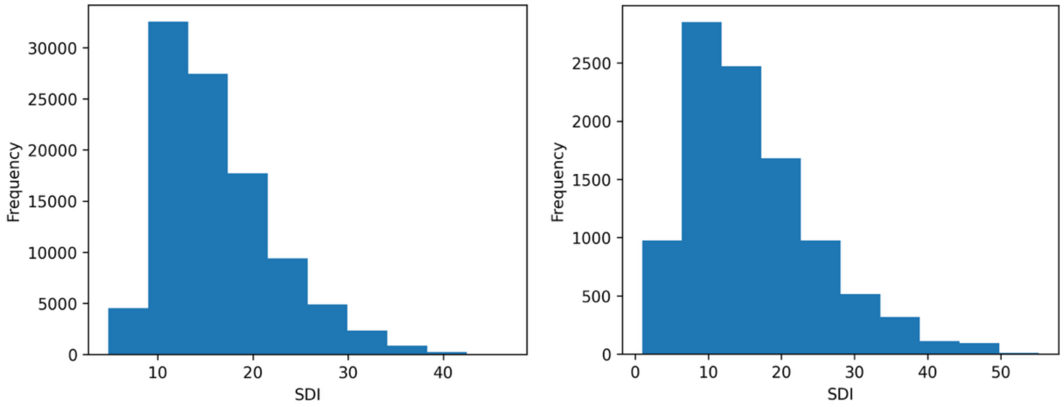


*(a)* *(b)*

*Figure S7: Comparison of census tract-level SDI distributions in (a) simulation cohort and (b) PRIME registry*
*SDI: social deprivation index*

### 6.10 Mortality adjustments by insurance status

In our secondary analysis, we further adjusted baseline age-sex-racial/ethnic-group-specific mortality rates by health insurance status, based on evidence from prior studies.[13,15] We assumed a constant race/ethnicity-specific prevalence of uninsured individuals and a hazard ratio for increased mortality risk among those uninsured. After age 65, we assumed all individuals received Medicare, and mortality rates returned to baseline values from the 2021 U.S. life tables.

### 6.11 Entering the health system

Our simulation made the assumption that individuals without health insurance entered the health system, defined as having a routine place for healthcare, more slowly than those with health insurance. We applied a hazard ratio ($HR_{\text{IHS}}$), where IHS refers to "in health system," to slow transition rates for simulated individuals without health insurance. The transition probability ($p_{IHS}$) and hazard ratio reduction ($HR_{\text{IHS}}$) were calibrated to match NHANES age-specific estimates of lacking a routine place for healthcare by insurance status. To obtain these NHANES estimates, we included 6,939 non-Hispanic Black and non-Hispanic white survey participants who were not currently treated with antihypertensive medication. In *Figure S8*, we compared the prevalence of the simulated cohort out of the health system (without a routine place of care) with NHANES age-specific estimates of having no place for routine healthcare by insurance status. The 95% confidence intervals for the NHANES age-specific estimates are shown as dots with error bars. Estimates for uninsured individuals after 65 are not included because the model assumes all individuals receive Medicare at age 65. Our simulation model reproduced the age-specific prevalence of not having routine access to healthcare in NHANES.

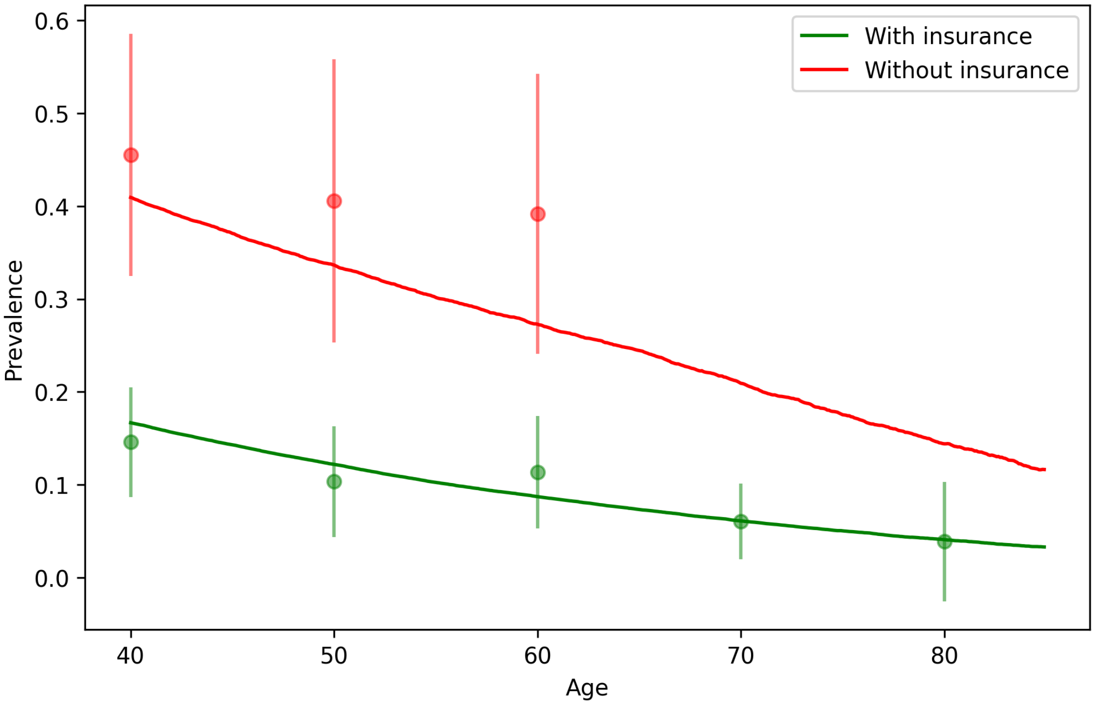


*Figure S8: Age-specific prevalence of not having a routine place for healthcare by insurance status in our simulation cohort (lines) and NHANES (dots)*
*NHANES: National Health and Nutrition Examination Survey*

### 6.12 Blood pressure adjustments out of the health system

We assumed that not having a routine place for healthcare affected systolic and diastolic blood pressure values. From NHANES, we selected participants aged at least 40 years who self-reported not currently taking antihypertensive medication (n=3,837 survey participants). Similar to our approach in *Section 6.4*, we estimated quantile regressions as a function of age, sex, no place for routine healthcare ($N_i$), and six SDI-related metrics ($S_i$) to predict systolic blood pressure values:

$$Y_i = \beta_0 + \beta_1 X_i + \beta_2 F_i + \beta_3 N_i + \sum_{j=1}^{k=6} \beta_j S_{ij},$$

and diastolic blood pressure values:

$$Z_i = \beta_0 + \beta_1 X_i + \beta_2 F_i + \beta_3 N_i + \sum_{j=1}^{k=6} \beta_j S_{ij}.$$

Because census tract-level SDI is not captured in NHANES, we included individual-level data on SDI components, including whether an individual was living in poverty, had less than 12 years of education, lived in a single-parent household, resided in rented housing, lived in overcrowded housing units, and was unemployed and younger than 65 years of age.[14] We then leveraged the estimated coefficients for the indicator of having no place for routine healthcare from each of the quantile regressions in our simulations (*Figure S9*). During simulation, if a simulated individual was not in the health system, the estimated coefficient was added to the individual's current systolic or diastolic blood pressure value while they were out of the health system.

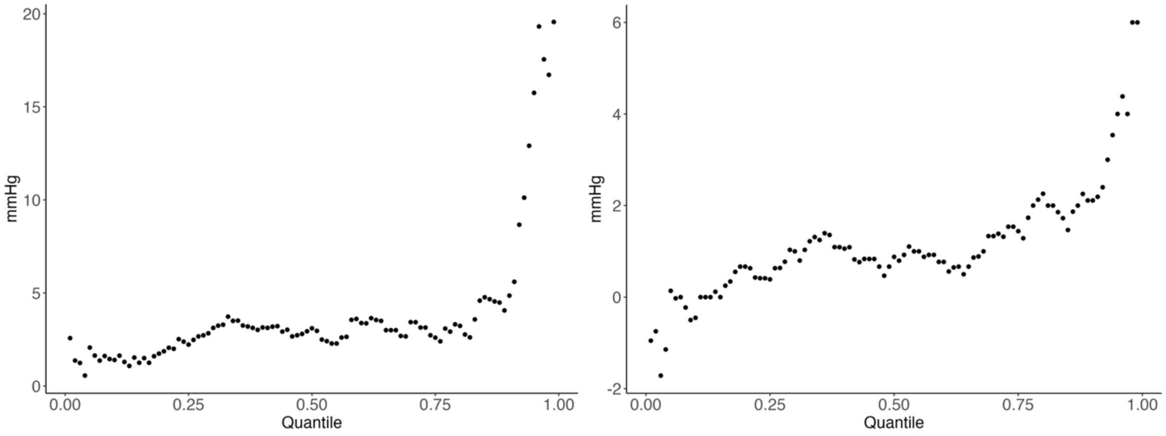


*(a)* *(b)*

*Figure S9: Blood pressure adjustments by quantile for not having a routine place for healthcare (a) systolic blood pressure and (b) diastolic blood pressure*

### 6.13 Additional results

*Table S10: Cumulative incidence of antihypertensive treatment by class under antihypertensive treatment guidelines with and without race adjustment*

| **Group** | **Race Adjustment** | | **No Race Adjustment** | |
|---|---|---|---|---|
| | Thiazide Diuretic | ACE Inhibitor | Thiazide Diuretic | ACE Inhibitor |
| Non-Hispanic Black | 76% | 0% | 30% | 46% |
| Non-Hispanic white | 30% | 45% | 30% | 45% |

*ACE inhibitor: angiotensin-converting enzyme inhibitor*

*Table S11: Main outcomes comparing antihypertensive treatment guidelines with and without race adjustment*

| **Outcome** | **Heterogeneous Treatment Effects** | | **No Heterogeneous Treatment Effects** | |
|---|---|---|---|---|
| | Race Adjustment | No Race Adjustment | Race Adjustment | No Race Adjustment |
| Cumulative incidence of hypertension stage 1 or 2 (%) | 90% [<1%] | 90% [<1%] | 90% [<1%] | 90% [<1%] |
| Years spent in stage 1 or 2 hypertension | 14.6 [<0.01] | 15.2 [<0.01] | 14.4 [<0.01] | 14.6 [<0.01] |
| Cumulative incidence of antihypertensive treatment | 76% [<1%] | 76% [<1%] | 76% [<1%] | 76% [<1%] |
| Years on antihypertensive treatment | 22.4 [<0.01] | 21.5 [<0.01] | 22.3 [<0.01] | 21.9 [<0.01] |
| Average age to initiate treatment | 52.4 [<0.01] | 52.4 [<0.01] | 52.4 [<0.01] | 52.4 [<0.01] |
| Years with uncontrolled hypertension | 8.1 [<0.01] | 8.6 [<0.01] | 7.8 [<0.01] | 7.9 [<0.01] |
| Years with controlled hypertension | 14.2 [<0.01] | 13.0 [<0.01] | 14.5 [<0.01] | 14.0 [<0.01] |
| Life expectancy | 38.1 [<0.01] | 37.9 [<0.01] | 38.1 [<0.01] | 38.1 [<0.01] |

*Mean [Monte Carlo standard error]*

### 6.14 Additional results for secondary analysis

*Table S12: Demographic and clinical characteristics of simulation cohort by race in secondary analysis*

| **Variable** | | **Non-Hispanic Black (N=50,000)** | **Non-Hispanic white (N=50,000)** |
|---|---|---|---|
| Female (%) | | 52% | 51% |
| With health insurance (%) | | 79% | 86% |
| With place for routine healthcare (%) | | 79% | 79% |
| Hypertension stage (%) | Normal | 29% | 38% |
| | Elevated | 15% | 15% |
| | Stage 1 | 36% | 34% |
| | Stage 2 | 19% | 14% |
| Census tract-level SDI (mean [SD]) | | 18.1 [6.4] | 14.6 [5.0] |
| Systolic blood pressure (mean [SD]) | | 124.1 [13.0] | 121.0 [12.7] |
| Diastolic blood pressure (mean [SD]) | | 78.5 [9.1] | 77.0 [9.1] |

*SD: standard deviation*

*Table S13: Main outcomes comparing antihypertensive treatment guidelines with and without race adjustment in secondary analysis*

| **Outcome** | **Heterogeneous Treatment Effects** | | **No Heterogeneous Treatment Effects** | |
|---|---|---|---|---|
| | Race Adjustment | No Race Adjustment | Race Adjustment | No Race Adjustment |
| Cumulative incidence of hypertension stage 1 or 2 (%) | 90% [<1%] | 90% [<1%] | 90% [<1%] | 90% [<1%] |
| Years spent in stage 1 or 2 hypertension | 15.6 [<0.01] | 16.1 [<0.01] | 15.4 [<0.01] | 15.6 [<0.01] |
| Cumulative incidence of antihypertensive treatment | 69% [<1%] | 69% [<1%] | 69% [<1%] | 69% [<1%] |
| Years on antihypertensive treatment | 21.3 [0.1] | 20.5 [0.1] | 21.3 [0.1] | 20.8 [0.1] |
| Average age to initiate treatment | 53.4 [<0.01] | 53.4 [<0.01] | 53.4 [<0.01] | 53.4 [<0.01] |
| Years with uncontrolled hypertension | 7.9 [<0.01] | 8.2 [<0.01] | 7.5 [<0.01] | 7.6 [<0.01] |
| Years with controlled hypertension | 13.4 [<0.01] | 12.3 [<0.01] | 13.7 [<0.01] | 13.1 [<0.01] |
| Life expectancy | 37.6 [<0.01] | 37.5 [<0.01] | 37.7 [<0.01] | 37.6 [<0.01] |

*Mean [Monte Carlo standard error]*

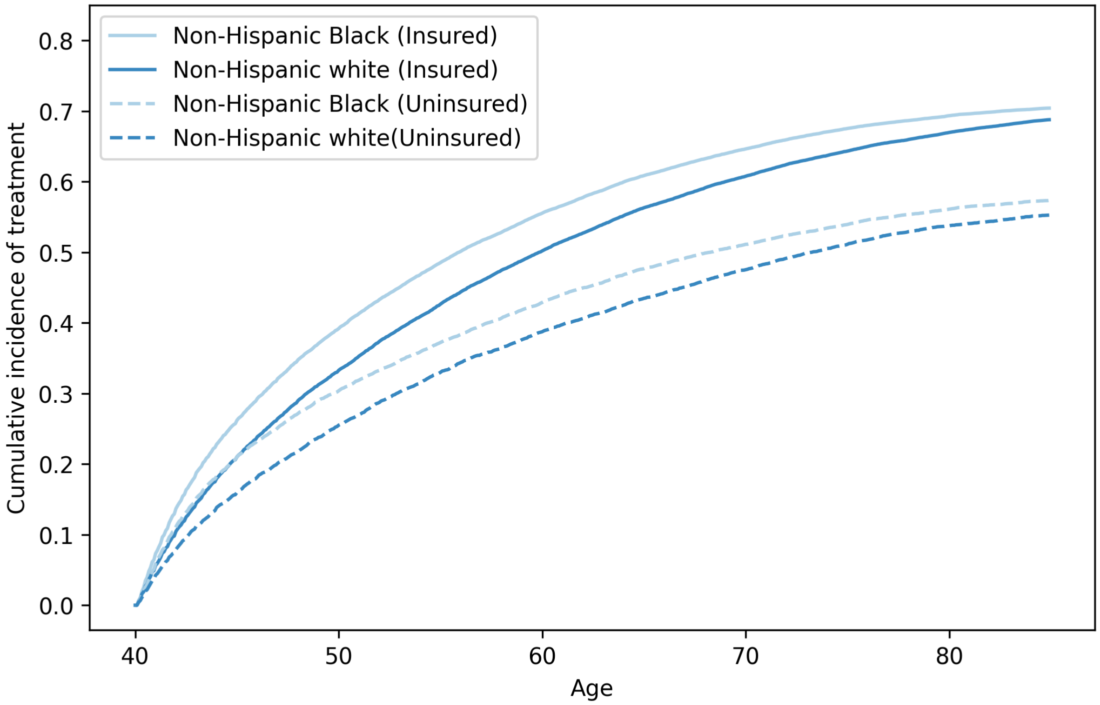


*Figure S10: Cumulative incidence of antihypertensive treatment for non-Hispanic Black and non-Hispanic white 40-year-old adults by insurance status*

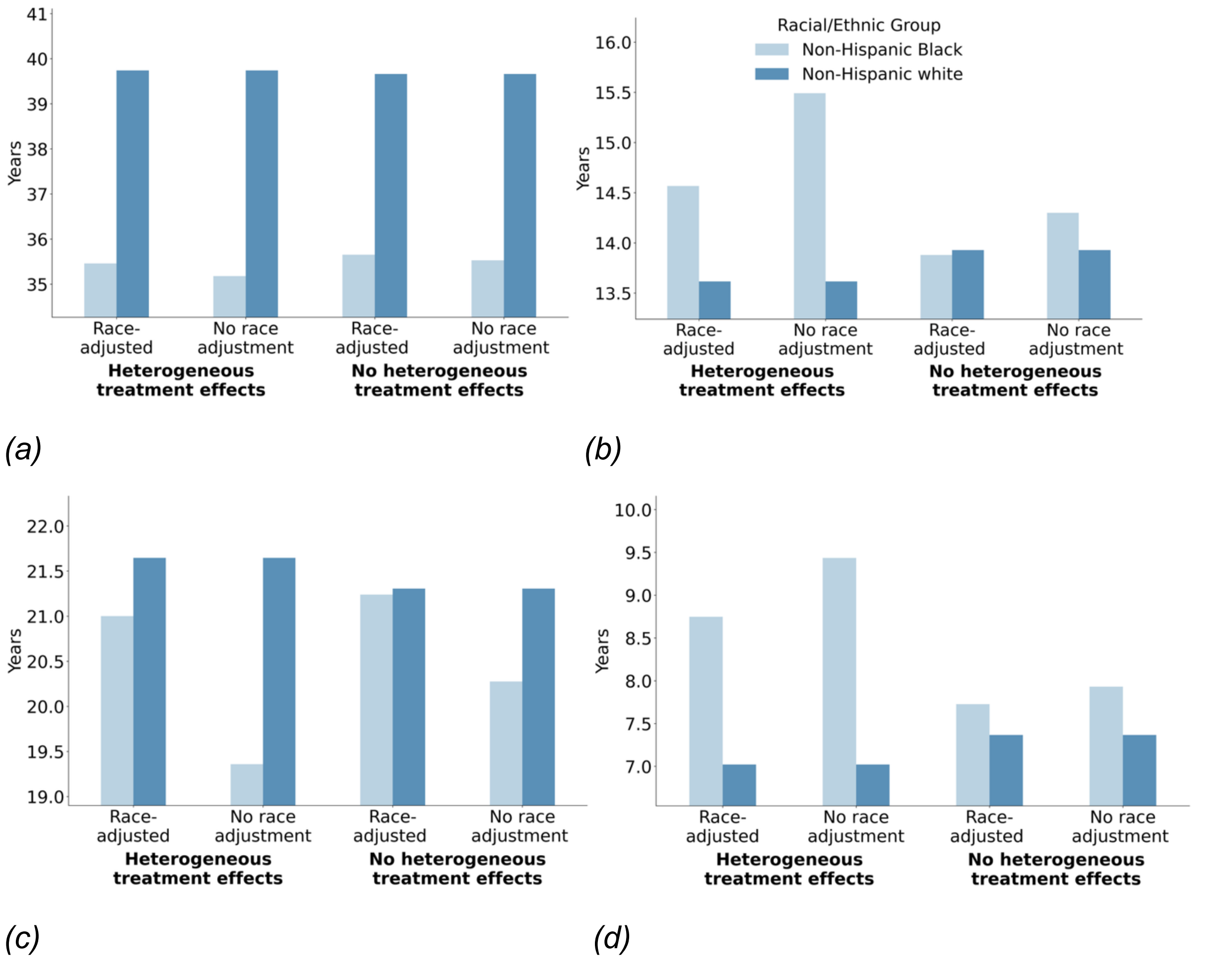


*Figure S11: Comparing antihypertensive treatment guidelines with and without race adjustment for non-Hispanic Black and non-Hispanic white adults (secondary analysis). Outcomes: (a) life expectancy, (b) years with stage 1 or 2 hypertension, (c) years on antihypertensive treatment, (d) years with uncontrolled hypertension*

*Table S14: Main outcomes for non-Hispanic Black and non-Hispanic white adults comparing antihypertensive treatment guidelines with and without race adjustment in secondary analysis*

| Outcome | Group | Heterogeneous Treatment Effects | | No Heterogeneous Treatment Effects | |
|---|---|---|---|---|---|
| | | Race Adjustment | No Race Adjustment | Race Adjustment | No Race Adjustment |
| Years spent in stage 1 or 2 hypertension | Non-Hispanic Black | 16.0 [0.1] | 17.1 [0.1] | 15.3 [0.1] | 15.7 [0.1] |
| | Non-Hispanic white | 15.2 [0.1] | 15.2 [0.1] | 15.5 [0.1] | 15.5 [0.1] |
| | Difference | -0.9 [0.1] | -1.9 [0.1] | 0.3 [0.1] | -0.2 [0.1] |
| Years on antihypertensive treatment | Non-Hispanic Black | 21.0 [0.1] | 19.4 [0.1] | 21.2 [0.1] | 20.3 [0.1] |
| | Non-Hispanic white | 21.6 [0.1] | 21.6 [0.1] | 21.3 [0.1] | 21.3 [0.1] |
| | Difference | 0.6 [0.1] | 2.3 [0.1] | 0.1 [0.1] | 1.0 [0.1] |
| Years with uncontrolled hypertension | Non-Hispanic Black | 8.7 [0.1] | 9.4 [0.1] | 7.7 [0.1] | 7.9 [0.1] |
| | Non-Hispanic white | 7.0 [<0.01] | 7.0 [<0.01] | 7.4 [0.1] | 7.4 [0.1] |
| | Difference | -1.7 [0.1] | -2.4 [0.1] | -0.4 [0.1] | -0.6 [0.1] |
| Life expectancy | Non-Hispanic Black | 35.5 [0.1] | 35.2 [0.1] | 35.7 [0.1] | 35.5 [0.1] |
| | Non-Hispanic white | 39.7 [0.1] | 39.7 [0.1] | 39.7 [0.1] | 39.7 [0.1] |
| | Difference | 4.3 [0.1] | 4.6 [0.1] | 4.0 [0.1] | 4.1 [0.1] |

*Mean [Monte Carlo standard error]*

*Table S15: Main outcomes for non-Hispanic Black and non-Hispanic white adults comparing antihypertensive treatment guidelines with and without race adjustment by insurance status in secondary analysis*

| **Outcome** | **Racial/Ethnic Group** | **Insurance** | **Heterogeneous Treatment Effects** | | **No Heterogeneous Treatment Effects** | |
|---|---|---|---|---|---|---|
| | | | Race Adjustment | No Race adjustment | Race Adjustment | No Race Adjustment |
| Years spent in stage 1 or 2 hypertension | Non-Hispanic Black | Yes | 15.6 [0.1] | 16.7 [0.1] | 14.7 [0.1] | 15.2 [0.1] |
| | | No | 17.8 [0.1] | 18.5 [0.1] | 17.2 [0.1] | 17.6 [0.1] |
| | Non-Hispanic white | Yes | 14.7 [0.1] | 14.7 [0.1] | 15.0 [0.1] | 15.0 [0.1] |
| | | No | 18.3 [0.2] | 18.3 [0.2] | 18.5 [0.2] | 18.5 [0.2] |
| Years on antihypertensive treatment | Non-Hispanic Black | Yes | 22.1 [0.1] | 20.5 [0.1] | 22.4 [0.1] | 21.4 [0.1] |
| | | No | 15.9 [0.2] | 14.2 [0.1] | 16.1 [0.2] | 15.1 [0.1] |
| | Non-Hispanic white | Yes | 22.3 [0.1] | 22.3 [0.1] | 22.0 [0.1] | 22.0 [0.1] |
| | | No | 16.4 [0.2] | 16.4 [0.2] | 16.0 [0.2] | 16.0 [0.2] |
| Years with uncontrolled hypertension | Non-Hispanic Black | Yes | 9.0 [0.1] | 9.8 [0.1] | 7.9 [0.1] | 8.2 [0.1] |
| | | No | 7.6 [0.1] | 7.9 [0.1] | 6.7 [0.1] | 6.8 [0.1] |
| | Non-Hispanic white | Yes | 7.1 [0.1] | 7.1 [0.1] | 7.5 [0.1] | 7.5 [0.1] |
| | | No | 6.3 [0.1] | 6.3 [0.1] | 6.6 [0.1] | 6.6 [0.1] |
| Life expectancy | Non-Hispanic Black | Yes | 36.1 [0.1] | 35.8 [0.1] | 36.3 [0.1] | 36.2 [0.1] |
| | | No | 33.0 [0.1] | 32.8 [0.1] | 33.1 [0.1] | 33.0 [0.1] |
| | Non-Hispanic white | Yes | 40.1 [0.1] | 40.1 [0.1] | 40.1 [0.1] | 40.1 [0.1] |
| | | No | 37.3 [0.2] | 37.3 [0.2] | 37.2 [0.2] | 37.2 [0.2] |

*Mean [Monte Carlo standard error]*
